# Prevalence and predictors of anemia in children under 5 years of age in sub-Saharan Africa: a systematic review and meta-analysis

**DOI:** 10.1101/2024.12.18.24319278

**Authors:** Ana Raquel Ernesto Manuel Gotine, Sancho Pedro Xavier, Melsequisete Daniel Vasco, Nerys Wendy Antonieta Alfane, Audêncio Victor

## Abstract

**Background:** Anemia is a significant public health challenge in Sub-Saharan Africa, particularly affecting children under five years old and posing serious health and developmental consequences. This systematic review and meta-analysis aim to quantify the prevalence and identify the predictors of anemia in this vulnerable age group across the region.

**Methods:** A systematic review and meta-analysis were conducted in adherence to PRISMA guidelines. Observational studies reporting on the prevalence and predictors of anemia among children under five in Sub-Saharan Africa were included. Comprehensive searches were performed in PubMed, Scopus, Web of Science, and Embase databases. The Joanna Briggs Institute (JBI) tools were used for critical appraisal. A random-effects model was applied to estimate pooled prevalence, while heterogeneity and publication bias were assessed using R software.

**Results:** A total of 32 studies comprising 93,388 children were included. The pooled prevalence of anemia was 55%, reflecting a widespread public health issue, with significant heterogeneity (I² = 99.7%). Key predictors included younger age (particularly 0–23 months), male gender, and indicators of poor nutritional status such as stunting. Socioeconomic and environmental factors, including rural residency and low maternal education, were also strongly associated with higher anemia rates. The observed variability across studies highlights the multifaceted nature of anemia’s determinants in the region.

**Conclusion:** The consistently high prevalence of anemia among children under five in Sub-Saharan Africa underscores the need for integrated public health strategies. Efforts to address nutritional deficiencies, improve maternal education, and enhance living conditions are essential to mitigate the burden of anemia and improve child health outcomes in the region.

## Introduction

Iron deficiency anemia is the most common nutritional disorder worldwide, affecting both developed and developing countries [1]. In the age groups of 0-23 months and 24-59 months, it is characterized by a hemoglobin concentration less than 105 g/L and 110 g/L, respectively [2,3]. According to the World Health Organization (WHO), about one-quarter of the global population, or approximately 1.6 billion people, are affected by this condition [4], with children and women of reproductive age being the most vulnerable groups [5]. Currently, it is estimated that about 40% of children between 6 and 59 months suffer from anemia worldwide [6], with Africa having the highest prevalence rates [4]. In Africa, more than 60% of children are anemic, and over 40% of these falls into the category of severe anemia [2,3]. In Sub-Saharan Africa, the prevalence of anemia among children under 5 varies from 42% in Eswatini to 91% in Burkina Faso [7].

The consequences of anemia during childhood include delayed growth, poor academic performance, disturbances in motor and cognitive development, increased vulnerability to infections, and increased morbidity and mortality[6]. Early acquired mental deficits are believed to be irreversible, and the consequences can persist even after treatment, emphasizing the importance of early detection and prevention[8].

The causes of anemia in this age group are complex and multifactorial. Among them are the child’s sex [1], low birth weight, undernutrition, poor socioeconomic status, household food insecurity, duration of breastfeeding, poor dietary iron intake, low maternal education level, diarrhea, fever, poverty, poor sanitation and hygiene conditions, monotonous diet, parental education level, and maternal anemia. These factors have been significant contributors to anemia in children under five years of age[9–11].

Despite significant efforts by African governments and various stakeholders to combat anemia, this condition still represents a critical public health challenge among children under five years old [12]. Studies conducted in different Sub-Saharan African countries have revealed variability in the predictors of anemia [1,13,14]. However, a comprehensive analysis that considers both the global prevalence and the predictors of anemia in this age group is still lacking. Understanding the dynamics of this condition is crucial to galvanize governmental action and resource mobilization, creating an enabling environment for the implementation of effective evidence-based interventions. Thus, this study aims to identify not only the prevalence of iron deficiency anemia among children under five years old in Sub-Saharan Africa but also the predictors for this condition.

## Materials and Methods

### Study design

This work consists of a systematic review and meta-analysis, aiming to assess the prevalence and predictors of anemia in children under five years old in Sub-Saharan Africa. The study follows the PRISMA (Preferred Reporting Items for Systematic Reviews and Meta-Analyses) guidelines, which ensure transparency and methodological rigor at all stages of the review and analysis process [15]. This study was registered in the international prospective register of systematic reviews (PROSPERO) under reference number CRD42024587000, available at: https://www.crd.york.ac.uk/prospero/display_record.php?ID=CRD42024587000

### Search Strategy

A comprehensive search was conducted in the electronic databases PubMed, Scopus, Web of Science, and Embase, with the aim of identifying relevant studies published up to the search date. The search strategy included combinations of keywords related to the topic, such as: PubMed (“Anemia” OR “Iron-deficiency anemia“) AND (“Predictors” OR “Determinants” OR “Risk factors” OR “Associated factors” OR “Causes“) AND (“Children under 5” OR “Infants” OR “Young children” OR “Preschool children“) AND (“Sub-Saharan Africa” OR “Africa south of the Sahara“). Scopus (iron-deficiency AND anemia) AND predictors OR (risk AND factors) AND (children AND under 5) OR infants OR (young AND children) OR (preschool AND children) AND (sub-saharan AND africa). Web of Science (“iron-deficiency anemia” AND predictors AND risk factors AND “children under 5” AND “sub-saharan africa” OR “africa south of the sahara“). Embase (’anemia’/exp OR ‘anemia’ OR ‘iron-deficiency anemia’/exp OR ‘iron-deficiency anemia’) AND (’predictors’/exp OR ‘determinants’ OR ‘risk factors’ OR ‘associated factors’ OR ‘causes’) AND (’children under 5’ OR ‘infants’/exp OR ‘young children’) AND (’sub-saharan africa’/exp OR ‘africa south of the sahara’).

### Inclusion and exclusion criteria

Observational studies (cross-sectional, cohort, or case-control) that reported the prevalence and predictor factors of iron deficiency anemia in children under 5 years of age in Sub-Saharan Africa were included in the review. To be included, studies had to meet the following criteria: Population: Children under 5 years located in countries of Sub-Saharan Africa. Exposure: Factors associated with anemia, including nutritional deficiencies, socioeconomic, and environmental conditions. Comparison: Children without anemia or with normal levels of hemoglobin. Outcome: Prevalence and severity of anemia, measured by hemoglobin concentration (hemoglobin levels <11g/dl) [2].Language: Studies published in English, Portuguese, or Spanish were included.

Studies not directly focused on children under five years old, as well as literature reviews and commentaries, were also excluded. Moreover, studies that did not study iron deficiency anemia and those that did not provide information about the tools used for diagnosing anemia were excluded. Studies limited to literature reviews, editorials, or conducted outside the context of Sub-Saharan Africa were also excluded. The identification, screening, and inclusion or exclusion of records process was carried out following the PRISMA flow diagram (Figure 1).

**Fig. 1.**
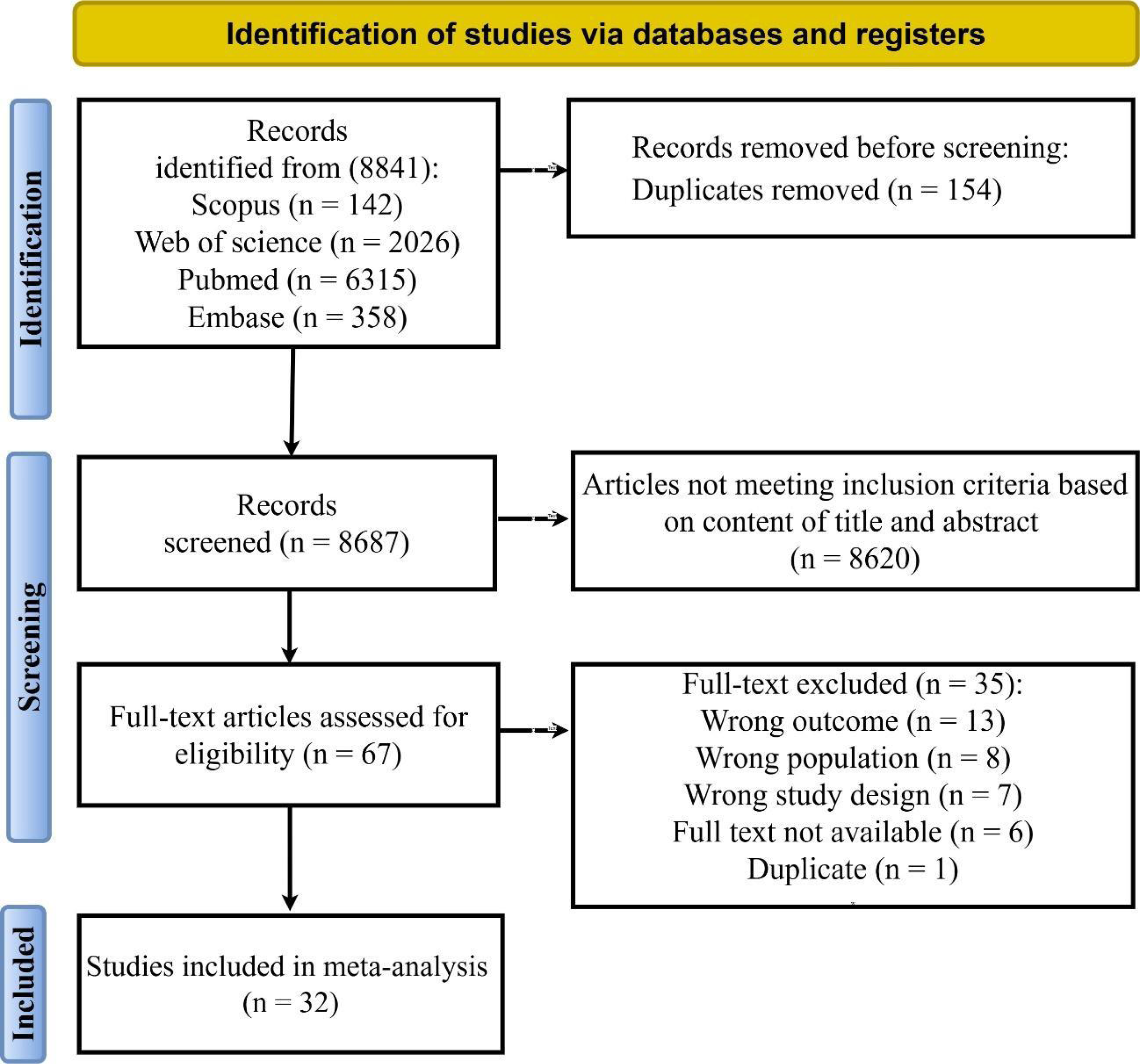
Flowchart of study selection.

### Study selection process

The selection of studies was conducted by two independent reviewers (AR and NW). Initially, all titles and abstracts of retrieved articles were screened to verify their relevance against the inclusion criteria. After the initial removal of duplicates, the reviewers independently examined all titles and abstracts, using the Rayyan tool [16]. Articles that were potentially eligible were then assessed in full. Any discrepancies between the reviewers were resolved by consensus, and, when necessary, a third reviewer (MDV) was consulted to resolve more complex issues. This process aimed to ensure the inclusion of only studies of high relevance and methodological rigor. The list of countries considered as part of Sub-Saharan Africa for this review includes: South Africa, Angola, Benin, Botswana, Burkina Faso, Burundi, Cape Verde, Cameroon, Chad, Comoros, Ivory Coast, Djibouti, Eritrea, Eswatini (Swaziland), Ethiopia, Gabon, Gambia, Ghana, Guinea, Guinea-Bissau, Equatorial Guinea, Lesotho, Liberia, Madagascar, Malawi, Mali, Mauritania, Mozambique, Namibia, Niger, Nigeria, Kenya, Central African Republic, Democratic Republic of the Congo, Republic of the Congo, Rwanda, São Tomé and Príncipe, Senegal, Sierra Leone, Seychelles, Somalia, South Sudan, Tanzania, Togo, Uganda, Zambia, and Zimbabwe.

### Data extraction

Data from the eligible studies were extracted by one reviewer (AREMG) into an Excel spreadsheet template suggested in a study [17,18],with a second and third reviewer (NWA and MDV) verifying each cell using a developed Excel spreadsheet. Information extracted included: author, year of publication, country where the study was conducted, study design, sample size, definition of anemia used in the study, and prevalence of anemia. After extraction, the data were verified by the three reviewers to ensure accuracy and consistency of the information collected.

### Quality assessment of studies (Risk of bias)

To individually assess the quality of the studies, protocols proposed by the Joanna Briggs Institute (JBI)[18] were used. For cross-sectional studies, the JBI critical appraisal checklist for analytical cross-sectional studies [19], was used, for longitudinal studies, the JBI critical appraisal checklist for cohort studies [20], and for case-control studies, the JBI critical appraisal checklist for case-control studies[21]. was employed. These are similar evaluative tools ranging from nine to eleven questions, used to assess the validity and reliability of the studies. Some of the checklist criteria include appropriate selection of cases and controls, precise measurement of exposures, control of confounding factors, and assessment of the temporality of exposures in relation to outcomes. The JBI checklists allow a systematic and standardized assessment of study quality, helping researchers identify potential sources of bias and interpret the results more reliably. For all types of designs, each item is rated as “yes,” “no,” “unclear,” or “not applicable (NA).” Questions with “NA” responses were not considered in the calculation. A percentage up to 49% indicates a low-quality study, a percentage from 50% to 70% indicates a moderate-quality study, and a percentage above 70% indicates a high-quality study [19].

### Statistical analysis

The meta-analysis was conducted to calculate the pooled estimates of anemia prevalence and the associated predictor factors. A random effects model (DerSimonian & Laird) was used due to the expected heterogeneity among the studies. The I² statistic and Q test were used to assess the heterogeneity among the studies [22,23]. Values of I² above 50% indicate substantial heterogeneity [23]. To investigate possible sources of this heterogeneity, subgroup analyses were conducted based on the year of publication (1994-2001, 2002-2009, 2010-2015, and 2016-2024), study design, and study quality (low, moderate, and high). The existence of publication bias was assessed by visual inspection of the symmetry of the funnel plot, and confirmed by Egger’s and Begg’s tests[24,25]. Values of p less than 0.05 indicated evidence of publication bias among the included studies. Results were presented in forest plots, showing point estimates with 95% confidence intervals (CI 95%). The Pooled Odds Ratio (OR) was used to describe the potential association between outcomes and predictors. A p-value of <0.05 was considered statistically significant.

### Sensitivity analysis

As heterogeneity assessments do not provide sufficient information about their sources, a meta-regression was conducted to explore potential factors responsible for the heterogeneity in the combined estimates. Additionally, a leave-one-out sensitivity analysis was conducted, iteratively removing one study at a time, to assess the impact of each study on the overall estimate. Further sensitivity analyses were conducted to test the robustness of the results, including the exclusion of low-quality studies or those with extreme estimates. A univariate meta-regression was also conducted to investigate the influence of design, quality, and year of publication of the studies [26].

## Results

### Characteristics of included studies

A total of 8,841 studies were identified in the following databases: 142 articles in Scopus, 2,026 in Web of Science, 6,315 in PubMed, and 358 in Embase. The flowchart details the process of selecting articles, highlighting reasons for exclusion. In the end, 32 articles were included for the Systematic Review and Meta-analysis (Figure 1). The 32 articles, published between the years 1995 and 2024, analyzed the prevalence and predictors of anemia in Sub-Saharan Africa. Among the included studies, 30 were cross-sectional, 1 was cohort, and 1 was case-control. The total sample of the review was 93,388 children between 0 and 59 months of age. The studies were conducted in the following countries: 7 in Ethiopia, 4 in Tanzania, 1 in Angola, 1 in Burundi, 1 in Cape Verde, 1 in Cameroon, 2 in Ghana, 2 in Guinea, 2 in Guinea-Bissau, 2 in Malawi, 2 in Namibia, 1 in Nigeria, 2 in Rwanda, 1 in Togo, 2 in Uganda, and 1 in Zambia **(Table 1).**

**Table 1:** Characteristics of all included studies.

| Authors | Year | Country | Study Design | Sample size | Mean/Median Age (months) | Age (months) | Outcome | Prevalence of Anemia |
| --- | --- | --- | --- | --- | --- | --- | --- | --- |
| Chao DL, Oron AP, Chabot CG, et al.[55] | 2022 | Nigeria | Cross-sectional | 11 536 | Not reported | 6–59 | Anemia | 67.9% |
| Premji Z, Yuna H, Clive S, et al.[56] | 1995 | Tanzânia | Cross-sectional | 338 | Not reported | 6 - 40 | Anemia | 74.1% |
| Tikua YS, et al.[57] | 2017 | Ethiopia; | Cross-sectional | 404 | 27.9 | 6–59 | Anemia | 51.4%. |
| Gebereselassie Y, et al.[58] | 2020 | Ethiopia | Cross-sectional | 422 | Not reported | 6–59 | Anemia | 41.7%. |
| Kobayashi E, Bharat N, Minato N, et al.[59] | 2018 | Zâmbia, | Cross-sectional | 4158 | Not reported | 6-59 | Anemia | 58%. |
| Shimanda PP, et al.[60] | 2019 | Namíbia | Cross-sectional | 1383 | 24 | 6–59 | Anemia | 49.6% |
| Angel MD, et al.[61] | 2017 | Rwanda | Cross-sectional | 650 | 34 | 6-59 | Anemia | 30.9% |
| Moschovis PP, Wiens MO, Arlington L, et al[47] | 2014 | Sudan | Cross-sectional | 163 | Not reported | 6-59 | Anemia | 80.4%. |
| Aheto JM K. \| Yakubu A, Adikwor E. Puplampu, et al.[62] | 2019 | Ghana. | Cross-sectional | 2434 | Not reported | 6–24 | Anemia | 54% |
| Sodde FM., Abebe D. Liga YN. Jabir, et al.[63] | 2023 | Ethiopia | Cross-sectional | 309 | 44.72 | 6-59 | Anemia | 51.7% |
| Barry TS, Oscar N, Nelson OO, Henry Mwambi.[64] | 2021 | Guinea | Cross-sectional | 2609 | Not reported | 0-59 | Anemia | 77% |
| Kajoba D, Walufu IE, Solomon M, et al.[65] | 2024 | Uganda | Cross-sectional | 364 | 14.1 | 6-23 | Anemia | 41.5% |
| Nkurunziza JC, Abukeera BN, Kalyango JN, et al.[66] | 2022 | Burundi | Cross-sectional | 501 | 15.24 | 6-24 | Anemia | 74.3% |
| Lweno O, Ellen Hamd, et al.[67] | 2022 | Tanzania | Cross-sectional | 2826 | Not reported | 6-12 | Anemia | 90% |
| Mamiro PS., Patrick K, Dominique R, et al.[68] | 2005 | Tanzania | Cross-sectional | 309 | Not reported | 3-23 | Anemia | 68% |
| Heinrichs H, Bilal SE, Tariku D, et al.[69] | 2020 | Ethiopia | Cross-sectional | 7324 | Not reported | 6-23 | Anemia | 72% |
| Oyedele O, et al.[70] | 2024 | Namibia | Cross-sectional | 2427 | Not reported | 0-59 | Anemia | 46.1% |
| Dyness K, Pammla M, Petrucka, HM, et al.[71] | 2018 | Tanzania | Cross-sectional | 436 | 20.3 | 6-59 | Anemia | 84.6% |
| Adish A, SA Esrey, TW Gyorkos, et al.[72] | 1998 | Ethiopia | Cross-sectional | 2080 | Not reported | 6-59 | Anemia | 42% |
| Shenton LM., Andrew D. Jones, Mark L. W, et al. [73] | 2020 | Ghana | Cross-sectional | 2388 | Not reported | 6–59 | Anemia | 40% |
| Malako BG., Melese ST, Tefera B, et al.[74] | 2018 | Ethiopia | Cross-sectional | 485 | 13.7 | 6–23 | Anemia | 52.6% |
| Ojoniyi OO., Clifford O. Odimegwu, Emmanuel O. [75] | 2019 | Tanzania | Cross-sectional | 7916 | Not reported | 0-59 | Anemia | 75.3 |
| Fançony C., Ânia S, João L, et al.[76] | 2020 | Angola | Cross-sectional | 948 | Not reported | 6 -23 | Anemia | 44.4% |
| Elmardi KA, Ishag A, Elfatih MM et al. [77] | 2020 | Sudan | Cross-sectional | 3094 | Not reported | 6–59 | Anemia | 49.4% |
| Menon MP, Steven S. Yoon[78] | 2015 | Uganda | Cross-sectional | 4065 | Not reported | 0–59 | Anemia | 9.6% |
| Nambiema A., Alexie R, Issifou Y et al. [49] | 2019 | Togo | Cross-sectional | 3378 | Not reported | 6 - 59 | Anemia | 70.9% |
| Endris BS., Geert JD, Seifu H, et al. [79] | 2022 | Ethiopia | Cross-sectional | 9267 | Not reported | 6 -59 | Anemia | 57% |
| Alamneh TS, Alemakef WM, Kassahun AG, et al. [80] | 2023 | Ethiopia | Cross-sectional | 8483 | Not reported | 6 - 59 | Anemia | 29.4% |
| Redd S.C., Jack J. W., Richard, W. S. [81] | 1994 | Malawi | Cohort | 252 | Not reported | 6 - 59 | Anemia | 25% |
| Kahigwa E, David Schellenberg, Sergi Sanz, John J. et al. [82] | 2002 | Tanzania | case–control | 216 | Not reported | 6–59 | Anemia | 55.6% |
| Woodruff BA., James PW, Ismael NT, et al. [83] | 2018 | Guinean | Cross-sectional | 5681 | Not reported | 0–59 | Anemia | 34.4% |
| Nkulikiyinka R., Agnes B., et al. [66] | 2015 | Rwanda | Cross-sectional | 4068 | Not reported | 6–59 | Anemia | 38.1% |

### Quality assessment of studies

The quality of the 32 studies included in this systematic review, comprising 30 cross-sectional studies, 1 longitudinal study, and 1 case-control study, was assessed using the critical appraisal tools from the Joanna Briggs Institute (JBI), specific to each study type. Of the 30 cross-sectional studies, 24 (73%) were classified as high quality, meeting at least 75% of the checklist criteria. These studies had well-defined inclusion criteria, reliable measurements, and appropriate statistical analyses. Nine studies (27%) were classified as moderate quality due to limitations in identifying and managing confounding factors, which could introduce bias into their findings. The single longitudinal study was rated as high quality, though it had some issues related to the completeness of follow-up of participants. The case-control study was rated as moderate quality due to incomplete comparability between cases and controls, as well as unclear periods of exposure measurement.

### Meta-analysis Prevalence of anemia

Figure 2 shows the prevalence of anemia derived from the 32 articles included in the overall analysis of this meta-analysis. The combined estimated proportion of anemia, obtained using the random effects model, was 0.55 (95% CI: 0.47; 0.63).

**Figure 2:**
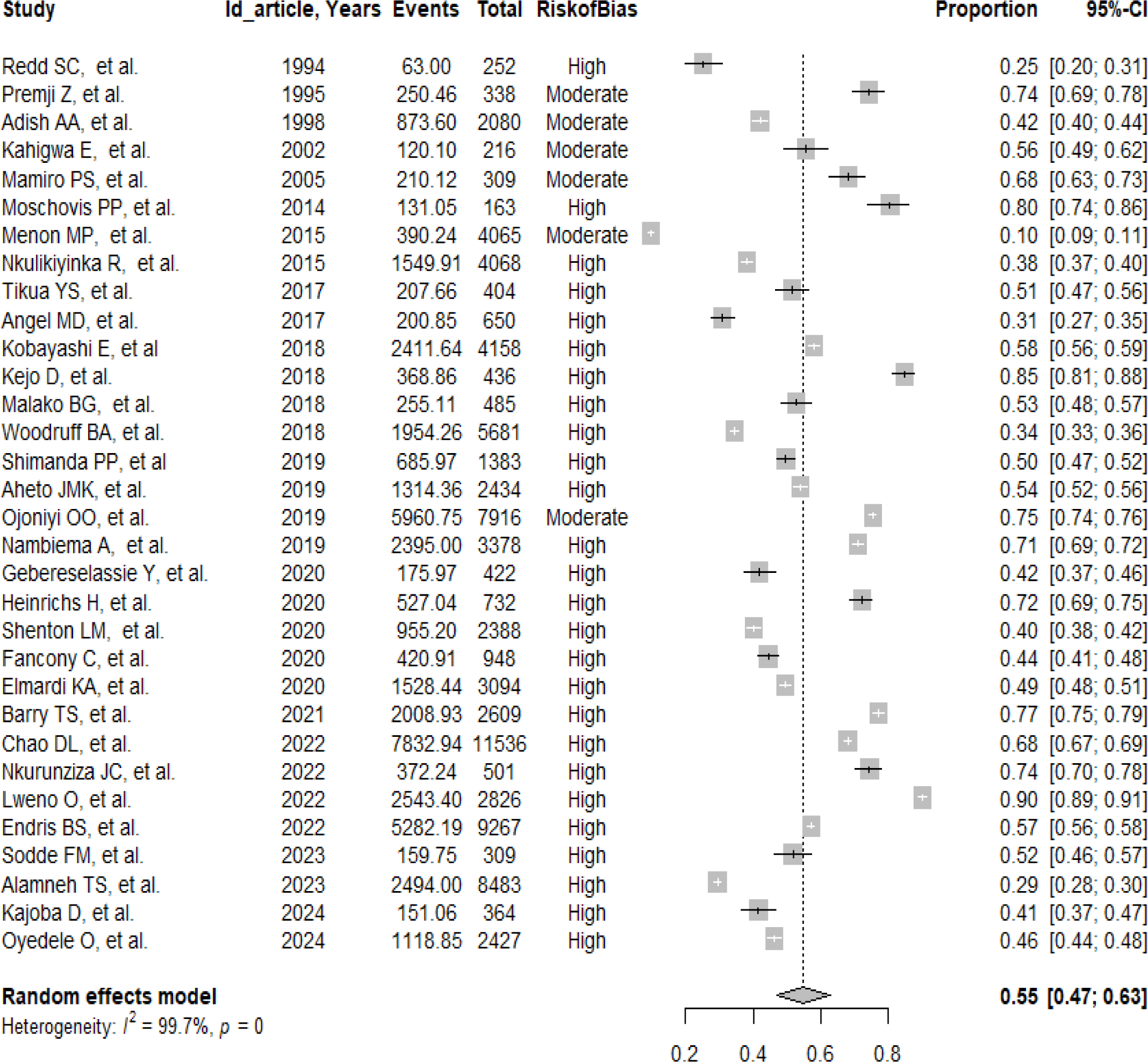
Prevalence of Anemia in Children Under 59 Months in Sub-Saharan Africa.

### Heterogeneity analysis

Figure 2 also presents the Forest Plot of the combined prevalence of anemia, indicating high heterogeneity among the studies. The I² index was 99.7% with a p-value < 0.001 in the Cochrane Q test, suggesting that the majority of the variability observed in the results is not explained by random errors, but by real differences among the studies (S2 – Figure 1).

### Subgroup analysis and meta-regression

Subgroup analyses were conducted based on publication year intervals (1994-2001, 2002-2009, 2010-2015, and 2016-2024), study design (case-control, cross-sectional, and cohort), and study quality (high, moderate, and low) **(Table 2)**. In the subgroup analysis stratified by publication year, the combined proportion of anemia varied over time. Between 1994-2001, the proportion was 0.47 (95% CI: 0.27; 0.68), with high heterogeneity (I² = 96.6%, p < 0.001). In studies published between 2002-2009, the proportion increased to 0.62 (95% CI: 0.36; 0.83), also with substantial heterogeneity (I² = 88%, p = 0.004). For the period 2010-2015, the proportion reduced to 0.38 (95% CI: 0.21; 0.60), while from 2016-2024, the proportion increased again to 0.57 (95% CI: 0.50; 0.64), both with I² > 99%, indicating great variation among the studies. Evaluating the study designs, the combined proportions also varied. In the cohort study, the proportion was 0.25 (95% CI: 0.20; 0.31), while the cross-sectional studies showed a higher combined proportion of 0.56 (95% CI: 0.49; 0.63), both with high heterogeneity (I² > 99%). The single case-control study presented a proportion similar to the cross-sectional studies at 0.56 (95% CI: 0.49; 0.62).

**Table 2:** Subgroup analysis and meta-regression for sources of heterogeneity.

| Characteristics | Number of studies | Proportion (CI95%) | I<sup>2</sup> (%) | p-value | Regression Coefficient (95% CI) | p-value |
| --- | --- | --- | --- | --- | --- | --- |
| **Publication year** |  |  |  |  |  |  |
| 1994-2001 | 3 | 0,47 (0,27; 0,68) | 96,6% | 0.001 | 1 |  |
| 2002-2009 | 2 | 0,62 (0,36; 0,83) | 88% | 0.004 | 0,61 (-0,76; 1,98) | 0,382 |
| 2010-2015 | 3 | 0,38 (0,21; 0,60) | 99,8% | 0.001 | -0,35 (-1,58; 0,87) | 0,571 |
| 2016-2024 | 24 | 0,57 (0,50; 0,64) | 99,7% | 0.001 | 0,41(-0,50; 1,33) | 0,377 |
| **Study design** |  |  |  |  |  |  |
| Case-control | 1 | 0,56 (0,49; 0,62) | 99,8% | 0.001 | 1 |  |
| Cross-sectional | 30 | 0,56 (0,49; 0,63) | 99,7% | 0.001 | 0,01 (-1,59; 1,61) | 0,989 |
| Cohort | 1 | 0,25 (0,20; 0,31) | 99,8% | 0.001 | -1,32 (-3,56; 0,91) | 0,246 |
| **Risk of bias** |  |  |  |  |  |  |
| High | 26 | 0,55 (0,48; 0,63) | 99,6% | 0.001 | 1 |  |
| Moderate | 6 | 0,52 (0,36; 0,68) | 99,9% | 0.001 | -0,13 (-0,86; 0,60) | 0,732 |
| **Total** | **32** | **0,55 (0,48; 0,62)** | **99,7%** | **0.001** |  |  |

Regarding the risk of bias, studies classified as high risk presented a combined proportion of 0.55 (95% CI: 0.48; 0.63) (I² = 99.6%, p < 0.001), while moderate-risk studies showed a slightly lower combined proportion of 0.52 (95% CI: 0.36; 0.68), still with substantial heterogeneity (I² = 99.9%, p < 0.001). Overall, the results indicate that the prevalence of anemia has been high in different periods, varying according to study type and risk of bias. The meta-regression analysis indicated that for anemia, the year of publication, study design, and study quality were not significant sources of heterogeneity **(Table 2).**

### Publication bias and sensitivity analysis

The funnel plot was used to assess publication bias in the meta-analysis of anemia prevalence. The plot (Figure 3) shows a relatively symmetric distribution of studies, suggesting an absence of publication bias. Begg’s test did not indicate significant bias (z = 0.62, p = 0.5377), reinforcing the notion that the distribution of studies is homogeneous in relation to the standard error, and Egger’s test also did not reveal significant bias (t = - 0.01, p = 0.9933), indicating a low probability of publication bias.

**Figure 3:**
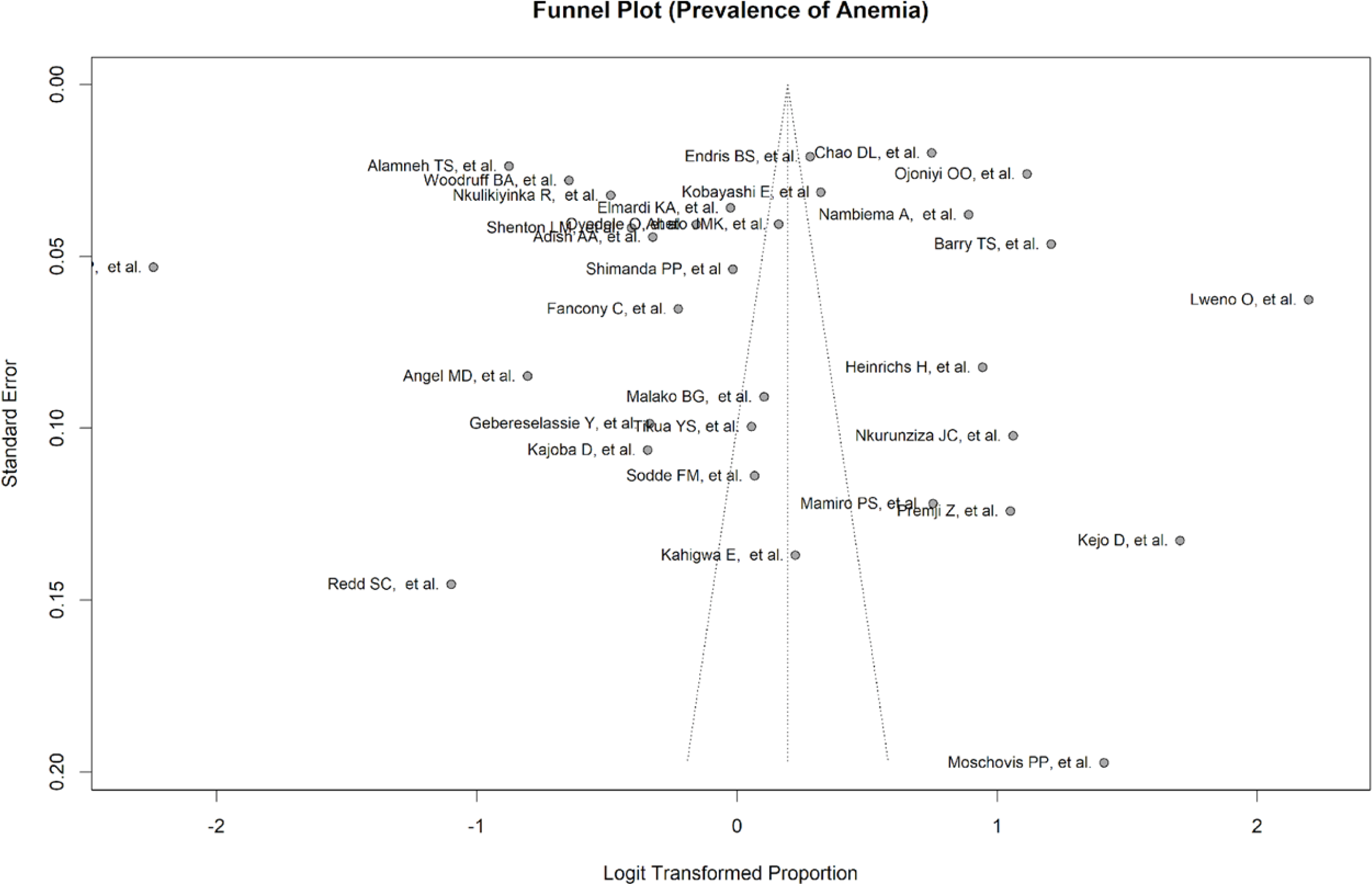
Publication bias of studies.

The sensitivity analysis (Figure 4) was performed to verify the impact of each study on the combined prevalence. The results demonstrate that the exclusion of any individual study did not significantly alter the combined prevalence, which remained stable at 0.55 (95% CI: 0.48; 0.62).

**Figure 4:**
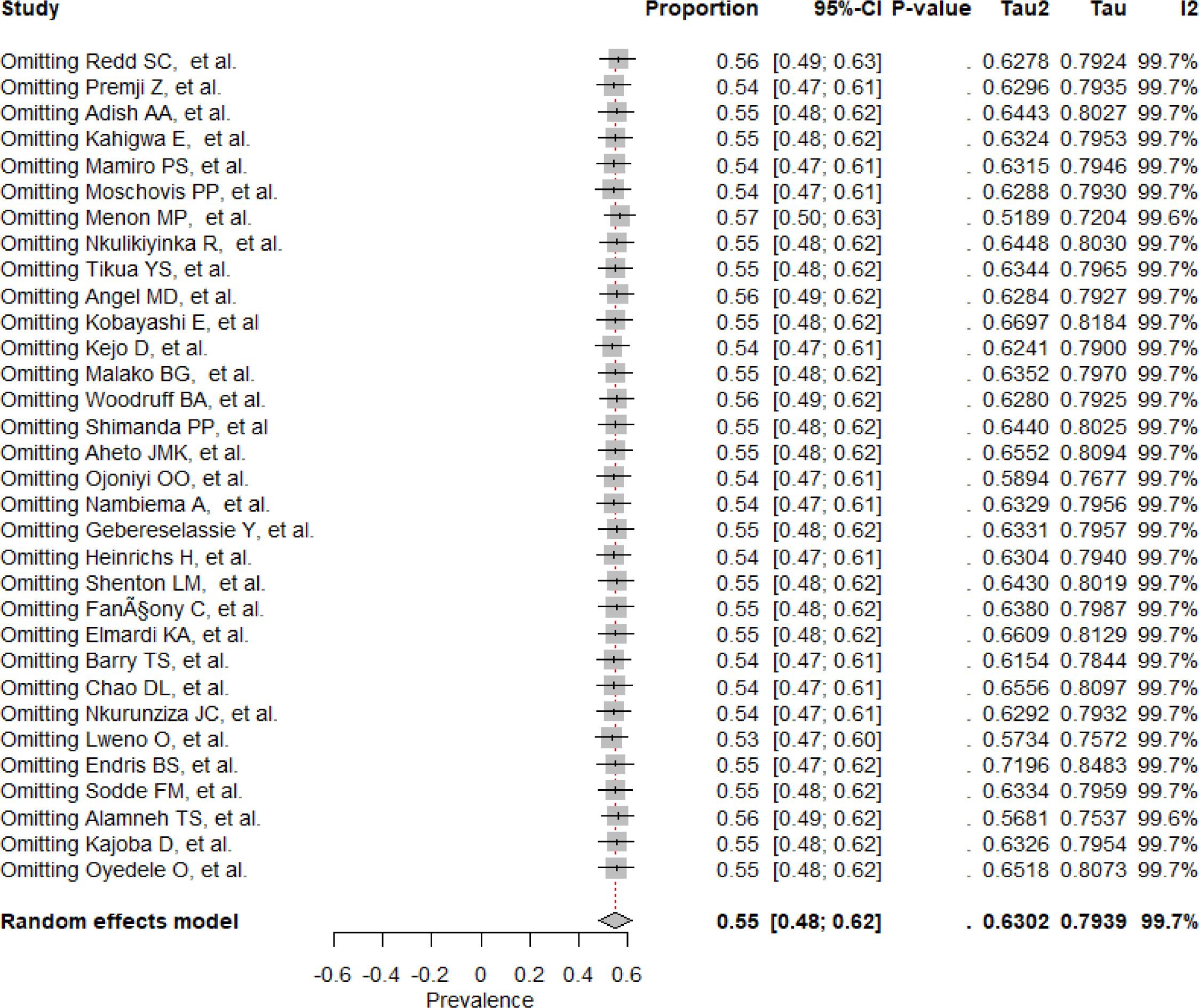
Sensitivity analysis.

### Predictors of Anemia Related to the child

The predictors of anemia in children under 5 years were: age between 0 and 23 months (OR = 1.60; 95% CI: 1.13–2.26), male gender (OR = 1.32; 95% CI: 1.10–1.60), malaria (OR = 3.18; 95% CI: 1.34–7.53), stunting (OR = 1.37; 95% CI: 1.12–1.67), fever (OR = 1.35; 95% CI: 1.05–1.74) **(S3 - Figure 2-15).**

### Related to the mother and household

Mothers aged 20 years or older (OR = 0.80; 95% CI: 0.65–0.99), mothers without formal education (OR = 1.33; 95% CI: 1.18–1.51), low dietary diversity (OR = 2.99; 95% CI: 2.01–4.46), residence in rural areas (OR = 1.20; 95% CI: 1.05–1.39), households with fewer than 5 people (OR = 1.37; 95% CI: 1.12–1.67), children belonging to the poorest economic group (OR = 1.46; 95% CI: 1.17–1.82) or the richest group (OR = 0.61; 95% CI: 0.46–0.82) (**S3 - Figure 16-25)**.

## Discussion

The combined prevalence of anemia in this age group was 55%. This prevalence was higher than reported in global studies [14], in Brazil [27] and in regions such as East Asia and Southeast Asia [28], but consistent with data from studies conducted in Africa [1], despite some differences in this region [28]. Although combined strategies, such as iron supplementation and management of infectious diseases including malaria, have been implemented by WHO to combat anemia [29], the condition remains a serious public health problem in Sub-Saharan Africa [30]. These results suggest that, at the current pace, achieving the global goal of reducing the prevalence of anemia by 50% by 2025 will be a significant challenge for the region [31].

In this study, various predictors were identified as associated with anemia in children. Among the child-related factors, age, gender, presence of malaria, stunting, and occurrence of fever stood out. Children aged between 0 and 23 months were at higher risk of having anemia compared to those older than 23 months. These results resemble findings from other studies [27,32]. This can be explained by the greater need for iron supplementation to allow for rapid growth and development, beyond insufficient intake. Moreover, as in this age group the immune system is not fully developed for enhanced defense and combating invasive agents, children are at higher risk of infections[33]. Additionally, it was evident in this study that most children with anemia, besides having malaria, also presented with fever. Malaria is well known to cause anemia [34–36], through two mechanisms: first, its high capacity to destroy red blood cells and second, by inducing anemia due to prolonged hypoferrimia (intestinal iron blockade)[14,27], after the release of hepcidin caused by systemic inflammation induced by the virus, triggering the liver to release the iron-regulating hormone, hepcidin [37]. Hepcidin effectively blocks iron absorption from the intestine [38], and this hepcidin-induced hypoferrimia can lead to iron deficiency anemia if prolonged [39]. Stunting was associated with the occurrence of anemia, with similar results found in other studies[13,40]. Stunting has been considered an indicator of chronic malnutrition, especially in children under five years [41,42]. Regarding child gender, in this study, boys were at higher risk of developing anemia, results consistent with findings in other research [13], although one study found no significant association [32]. This association may be explained by the fact that male children have a higher iron demand due to a faster growth rate [43].

Regarding predictors related to the mother and household, maternal age of 20 years or older was associated with a lower chance of anemia in children compared to teenage mothers. This shows that physical and emotional maturity, as well as a higher likelihood of socioeconomic stability, contribute to better health outcomes for the children of these mothers as observed in some studies [44,45]. Additionally, advanced maternal age may be associated with greater experience and knowledge about health and nutrition practices, contributing to better childcare [29,46,47]. Lack of formal education among mothers has been strongly linked to worse child outcomes [47]. Our findings showed that mothers without formal education have a significantly higher probability of having children with anemia. This resembles other studies conducted in Ethiopia and Togo that showed maternal education is a crucial factor influencing health and nutrition practices, thus impacting child health [48,49].

Results from this study showed that low dietary diversity is associated with a substantial increase in the risk of child anemia. This corroborates studies conducted in the Sub-Saharan region of Africa where low dietary diversity was one of the most significant factors associated with child anemia [50,51]. Dietary diversity is essential to ensure adequate intake of micronutrients, such as iron and vitamins, which are critical to preventing anemia[48]. Children residing in rural areas are more likely to be anemic. Limited access to healthcare services, lower availability of nutritious foods, and unfavorable socioeconomic conditions are factors contributing to this increased risk [47,52,53]. Unexpectedly, the presence of fewer than five people in the household was associated with an increase in the prevalence of anemia. This may likely be related to a smaller support network and limited resources for childcare [29,47]. Contrary to children of mothers belonging to wealthier economic groups, children belonging to poorer economic groups are more likely to develop anemia. This finding resembles other studies that showed poverty is linked to a range of negative outcomes, including food insecurity, limited access to healthcare services, and inadequate living conditions, factors that contribute to anemia [29,49]. Since wealth is associated with better living conditions, greater access to quality healthcare, and better nutrition, all factors that contribute to better health outcomes [47,54], the design and implementation of policies aimed at improving the economic situation of families can have a significant positive impact on children’s health. Meanwhile, children from wealthier economic groups are less likely to be anemic. Wealth is associated with better living conditions, greater access to quality healthcare, and better nutrition, all factors that contribute to better health outcomes [47,54]. However, it is important to consider that wealth alone does not guarantee the absence of health problems, but it offers a significant advantage in terms of resources available to prevent and treat adverse conditions. One of the main strengths of this study was the conduct of an exhaustive search, using various databases and comprehensive search strategies. However, a significant limitation was the presence of substantial heterogeneity among the included studies. Despite this limitation, most of the studies included demonstrated high methodological quality (73%), which reinforces the reliability of the results. These findings can contribute significantly to the formulation of more effective and evidence-based health policies.

### Conclusion

The prevalence of anemia in children under 5 years in Sub-Saharan Africa remains high, and the predictors of anemia identified in this meta-analysis highlight the importance of biological, nutritional, and socioeconomic determinants. Age between 0 and 23 months, male gender, low dietary diversity, presence of malaria, stunting, and fever were associated with a higher risk of anemia, reflecting frequent nutritional and health vulnerabilities in this age group. Moreover, contextual factors such as residence in rural areas, households with fewer than five people, mothers without formal education, and belonging to poorer economic groups reinforce the influence of the environment and socioeconomic conditions in the development of anemia. On the other hand, children whose mothers are aged 20 years or older and those from wealthier economic groups showed a lower risk, highlighting possible protective factors. These findings emphasize the need for integrated interventions that address dietary diversity, disease prevention such as malaria, and socioeconomic inequalities to reduce the prevalence of anemia in this age group.

## Data Availability

Data from the articles included in this systematic review and meta-analysis are available if needed and we are available to provide them.

## Acknowledgements

Not applicable.

## Authors’ Contributions

All authors agreed to be accountable for all aspects of the work and participated in all stages from the conception of the study idea, article searching, article quality assessment, data extraction, analysis, and interpretation. They also contributed to the drafting and revision of the manuscript. Based on this, the research idea was conceptualized by (AREMG). The three authors also independently and meticulously assessed the content of each of the identified articles (AREMG, NA, MDV). The three reviewers (AREMG, NA, MDV) conducted the study selection independently and blindly after a thorough screening of the abstracts and full texts of the studies. Data were extracted by (AREMG, NA, MDV, AV, and SX). The quality of the included studies was independently assessed by three authors (AV and SX). The data analysis and interpretation were conducted by (SX and AREMG). Finally, all authors gave their final review and approval.

## Declarations

Since the study did not use primary data, ethical approval is not applicable.

## Consent for Publication

Not applicable.

## Supporting information

S1 Table of quality assessment of the cross-sectional studies (n=30), using the JBI checklist, included in the systematic review

S1.1. Table of quality assessment of the longitudinal studies (n=1), using the JBI checklist, included in the systematic review:

S1.2. Table of quality assessment of the case-control studies (n=1), using the JBI checklist, included in the systematic review

S2 Figure1. Heterogeneity.

S3 Figure 2-25. Factors associated with anemia S4 File Prisma checklist.

**S 2:** Heterogeneity

**Figura 1:**
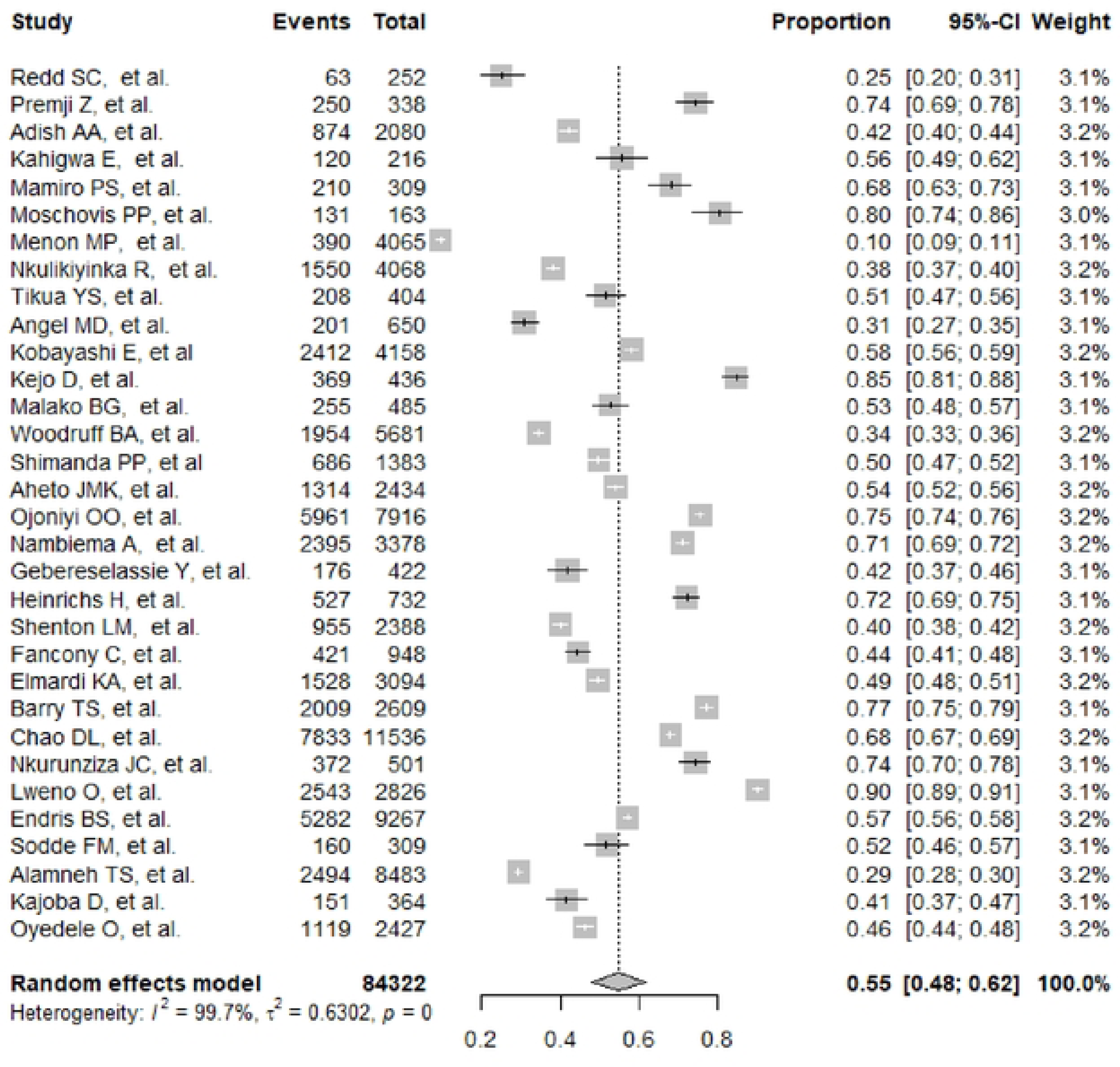
Heterogeneity.

**S 3:** Factors associated with anemia

**Figura 2:**
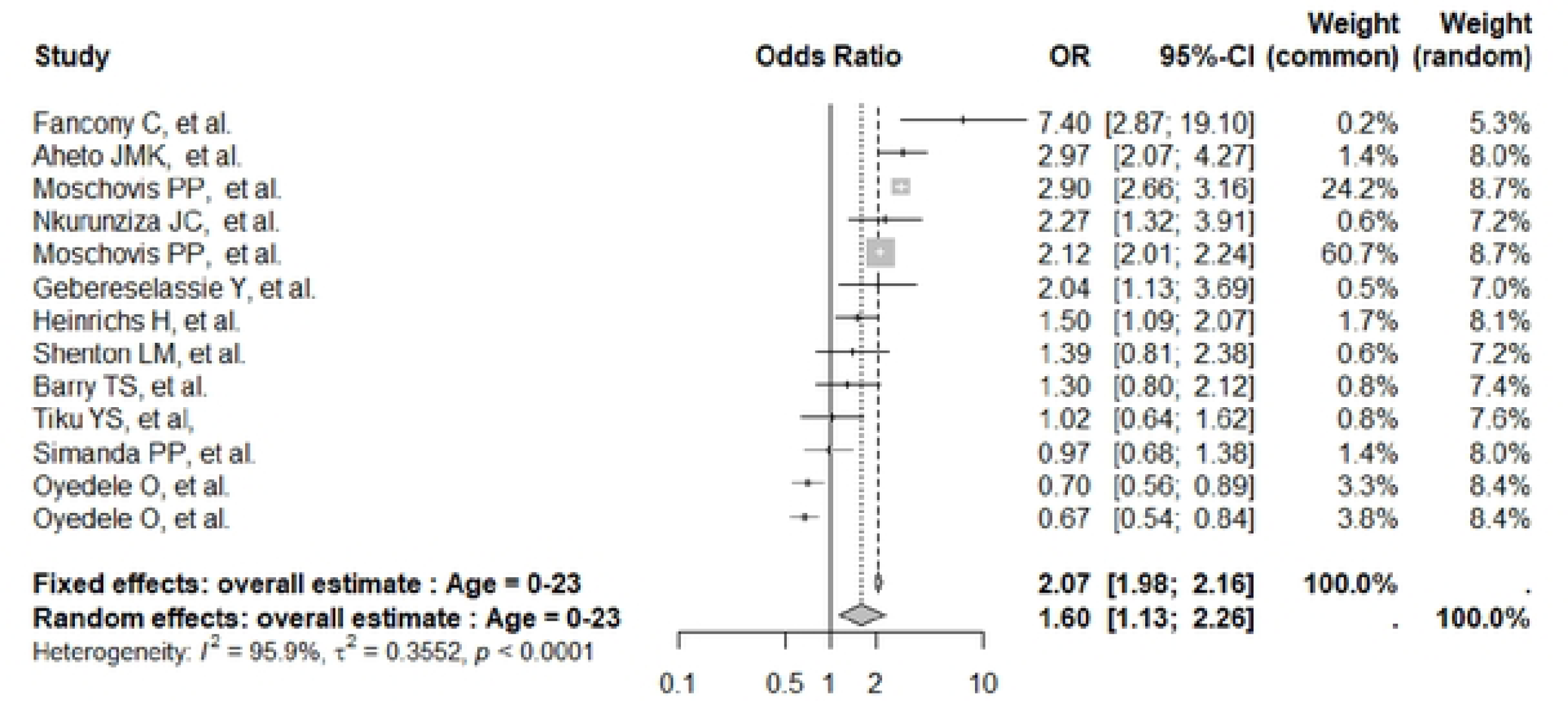
Child’s age O - 231nonths.

**Figura 3:**
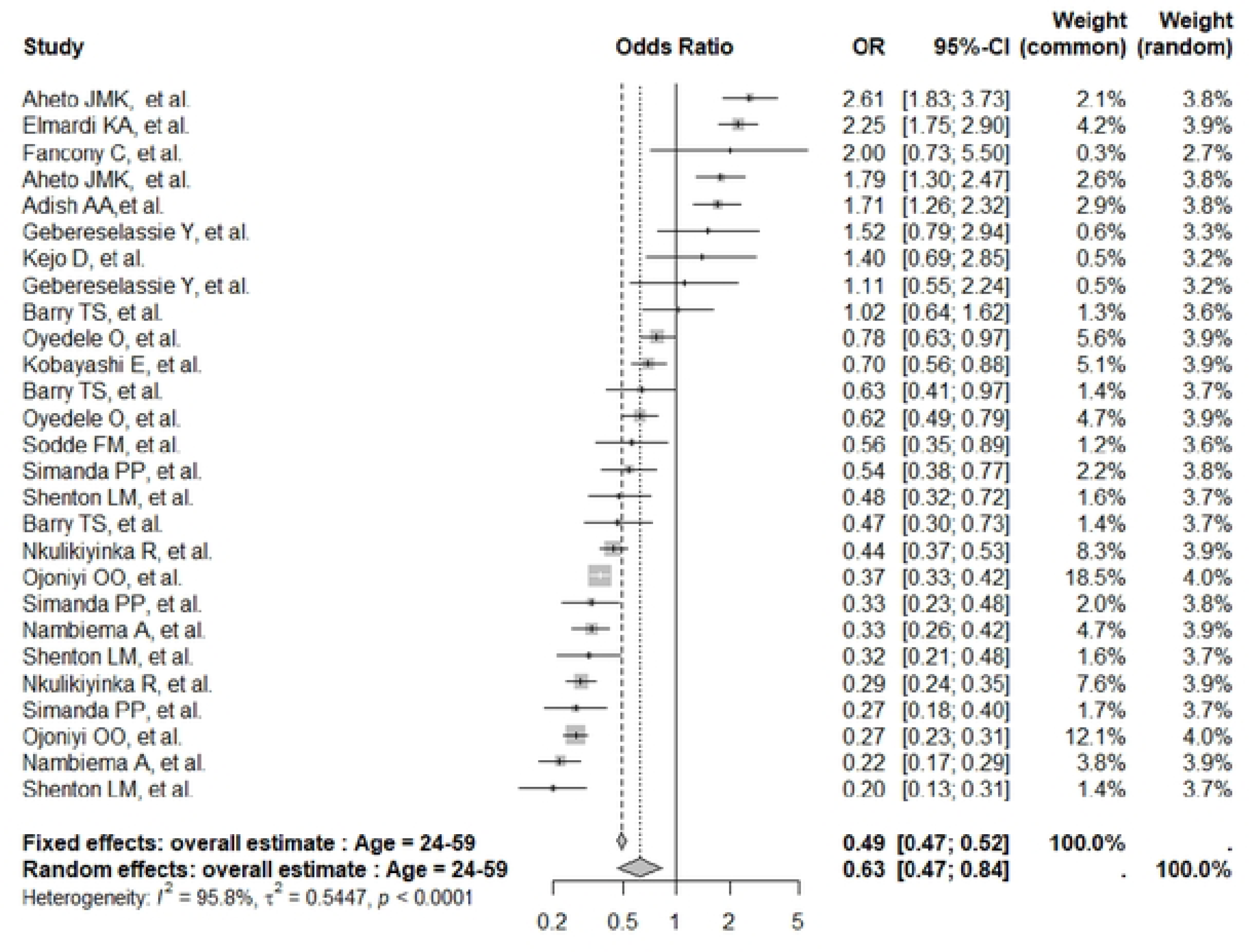
Child’s age 24 - 59 months.

**Figura 4:**
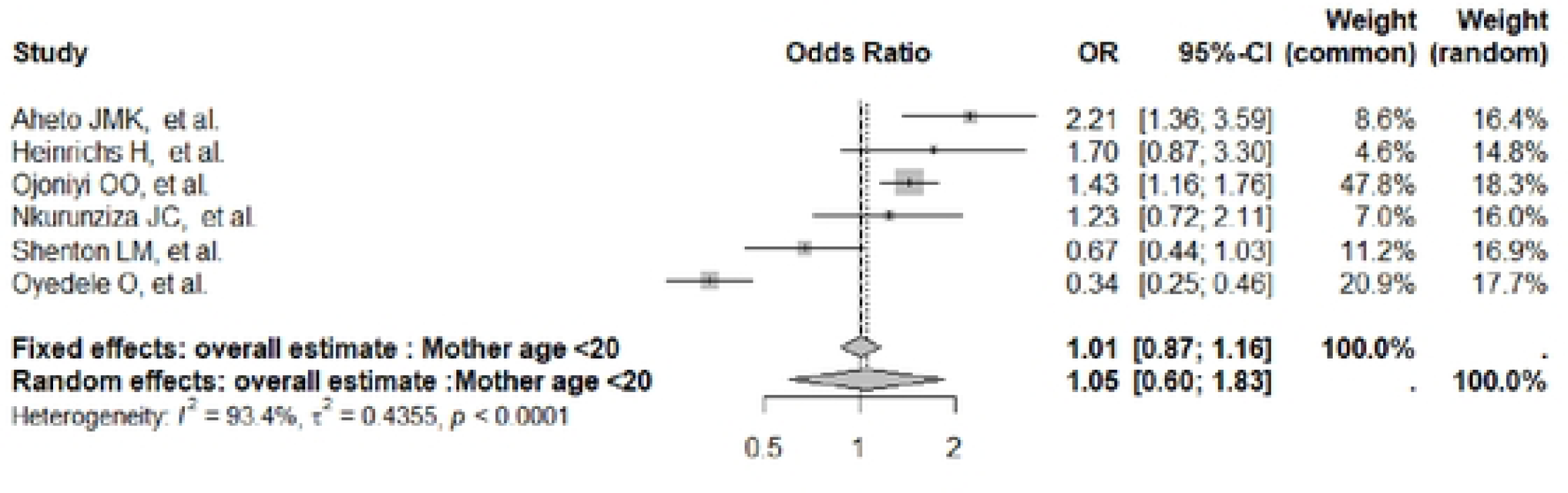
Mother’s age< 20 years.

**Figura 5:**
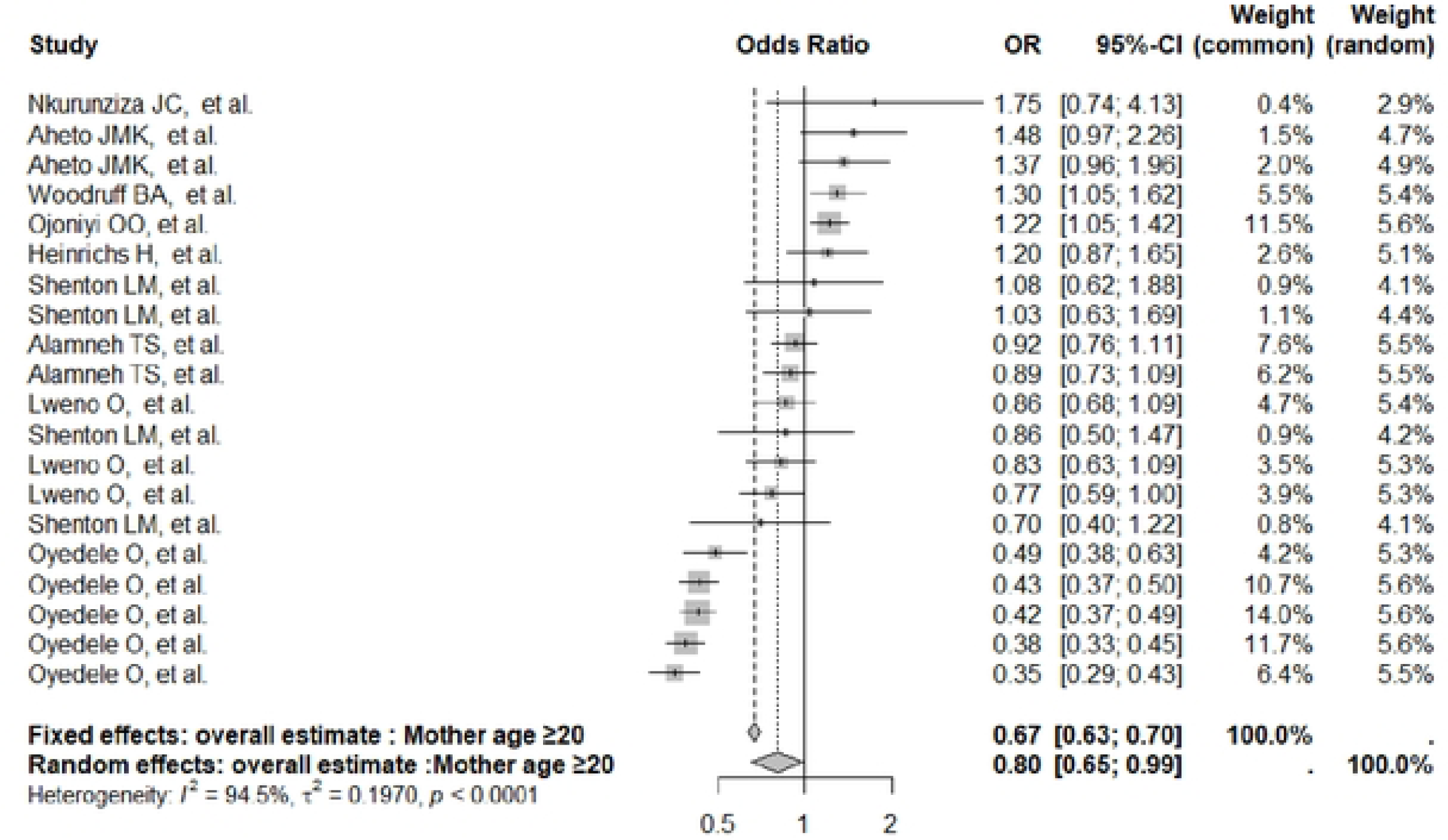
Mother’s age> 20 years.

**Figura 6:**
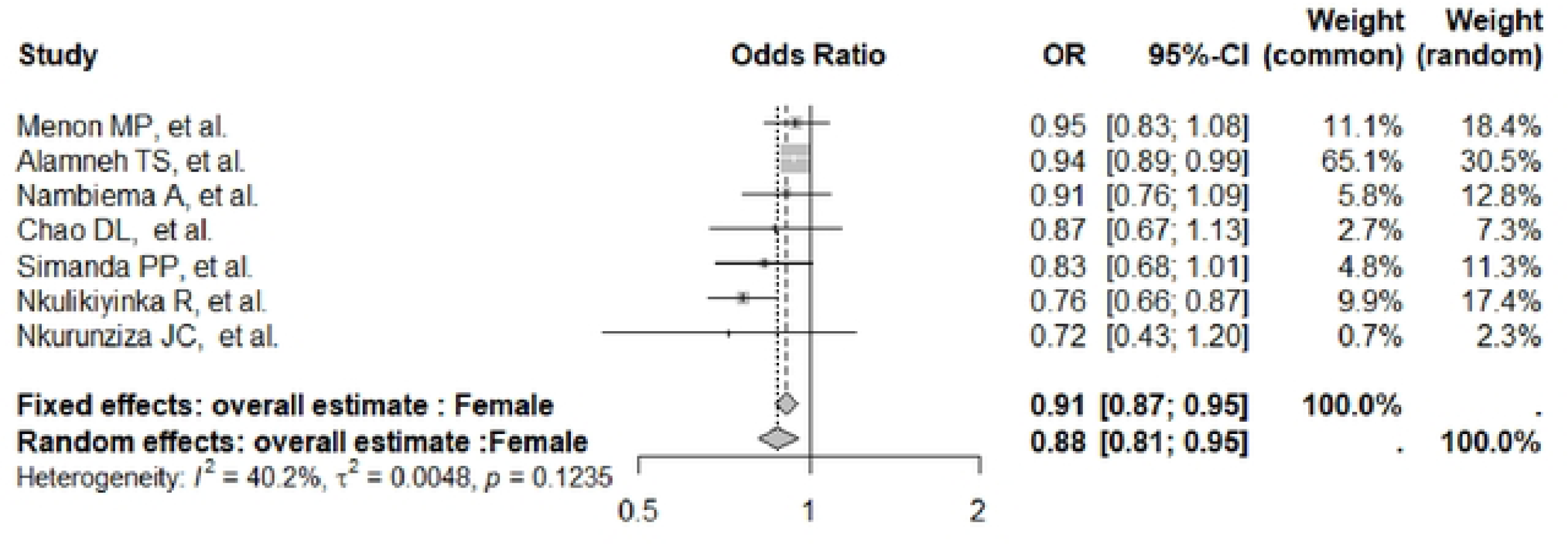
Child’s sex (female).

**Figura 6:**
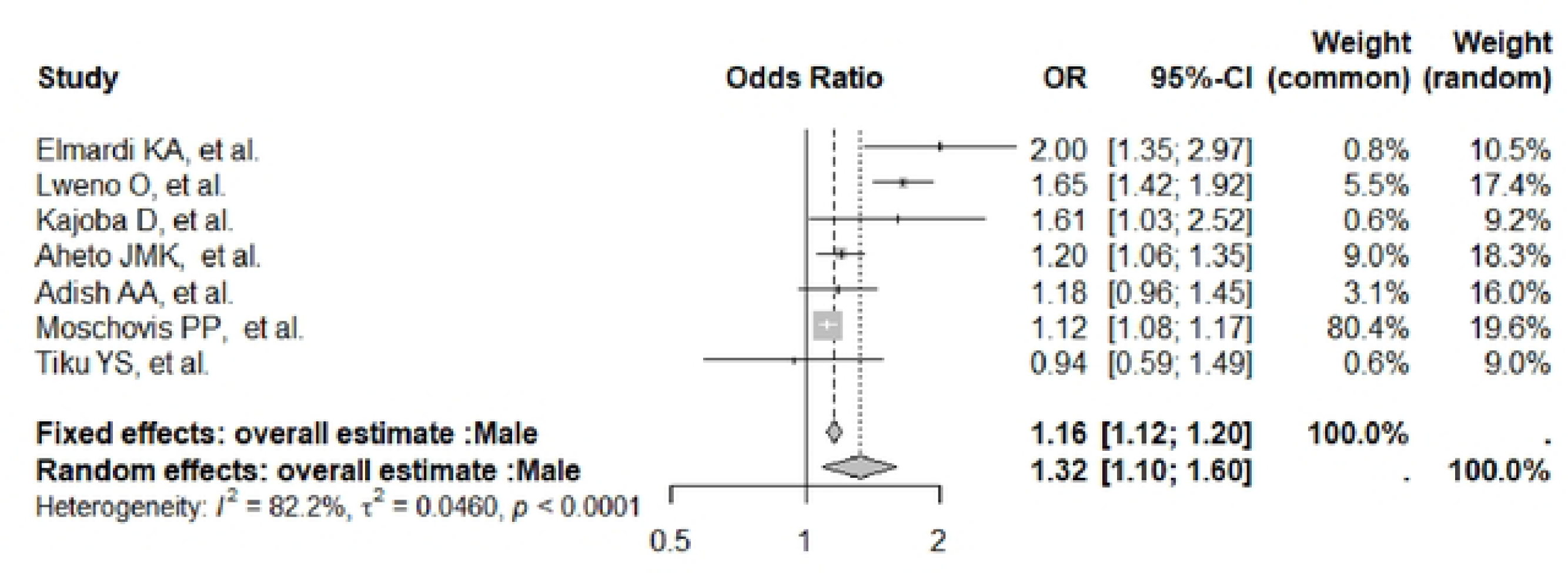
Child’s sex (male).

**Figura 7:**
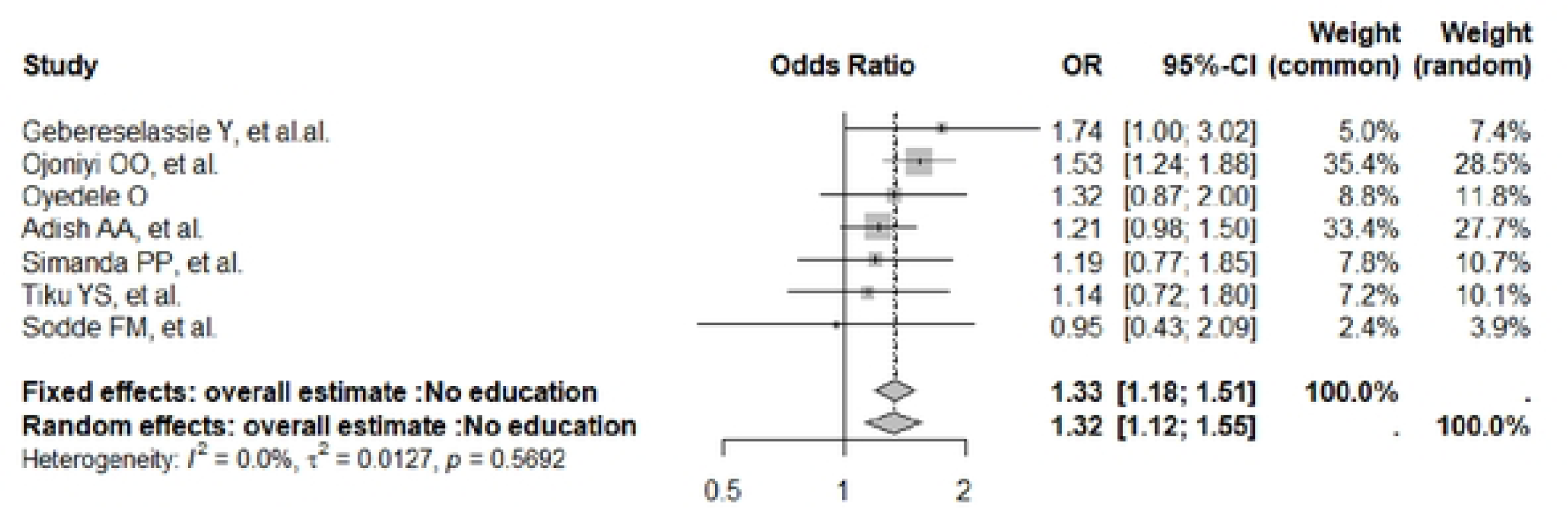
Mother education (no education).

**Figura 8:**
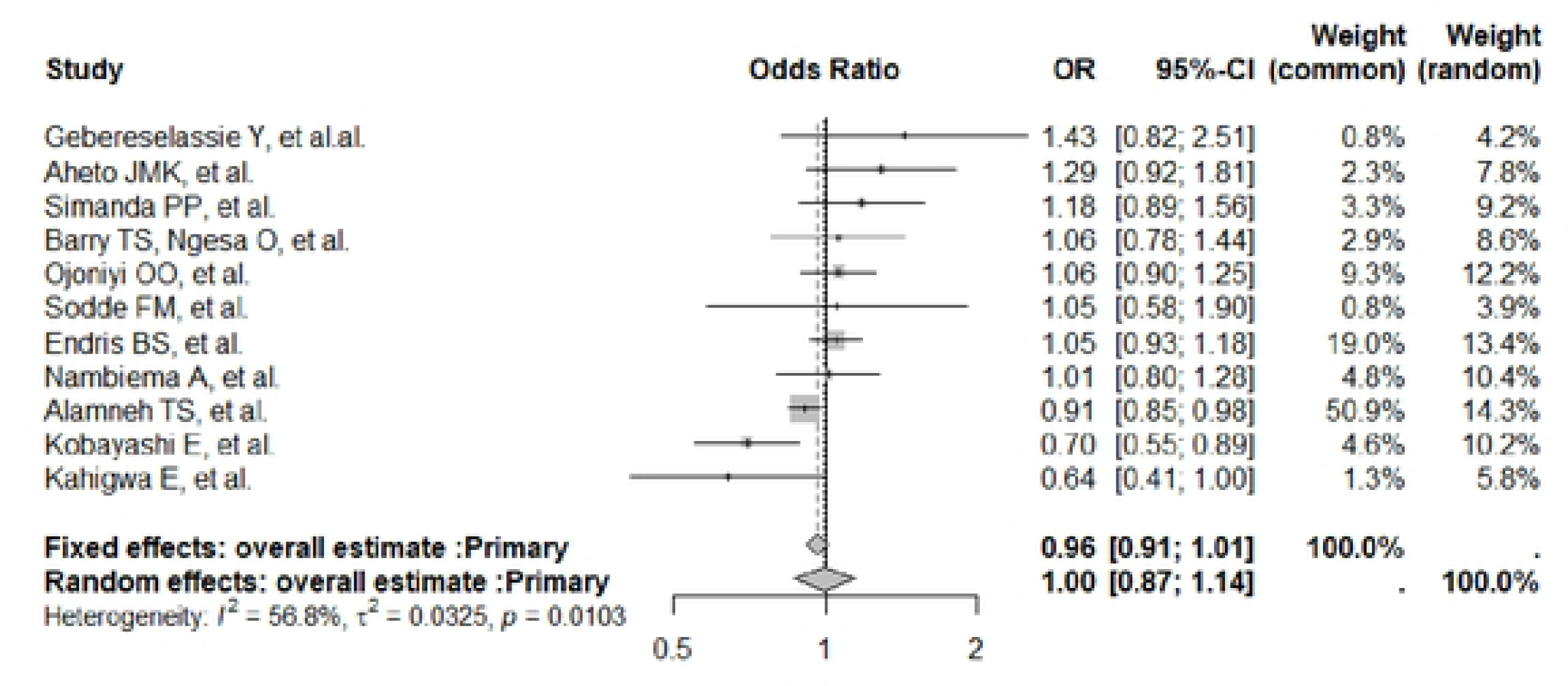
Mother education (primary).

**Figura 9:**
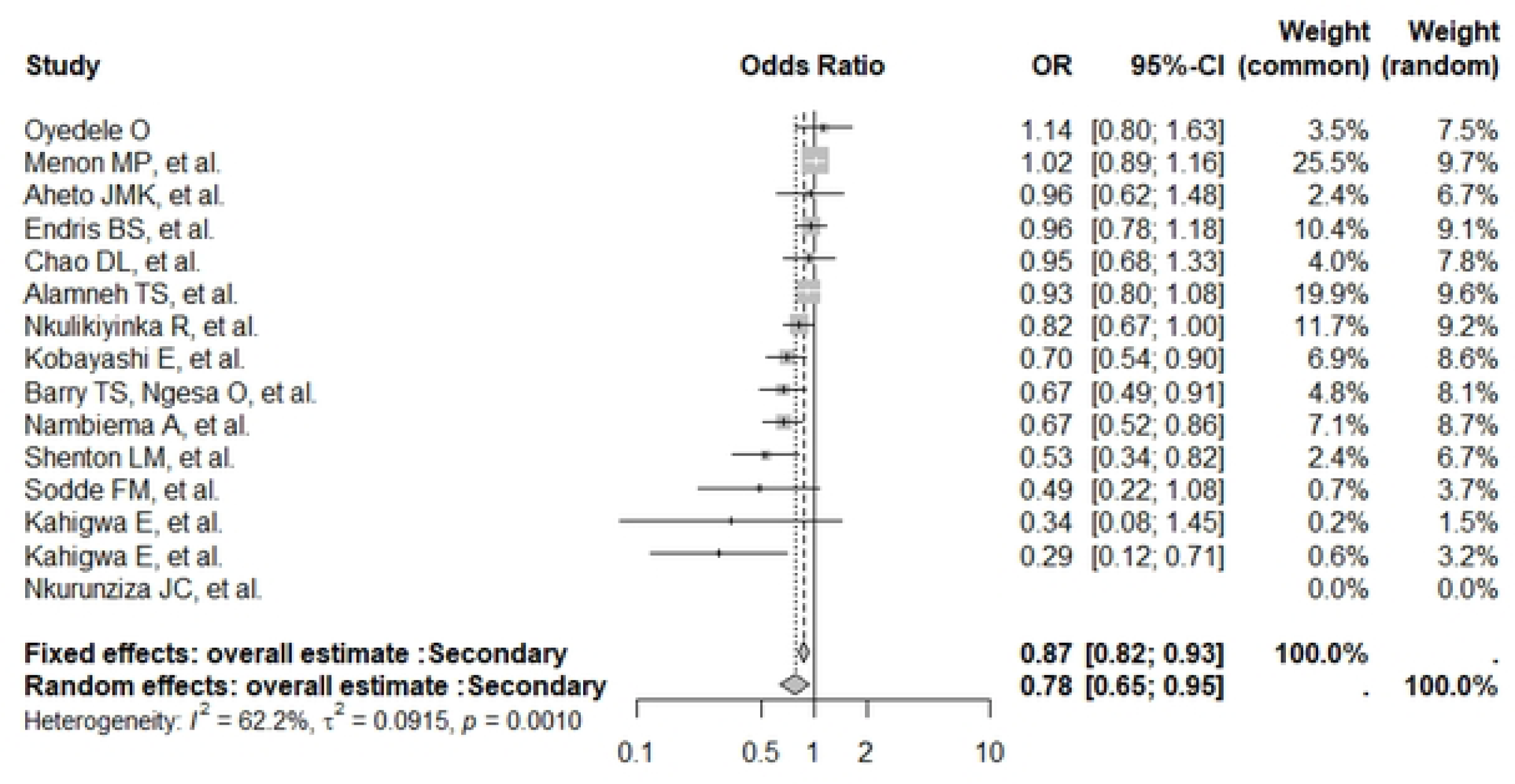
Mother education (secondary).

**Figura 10:**
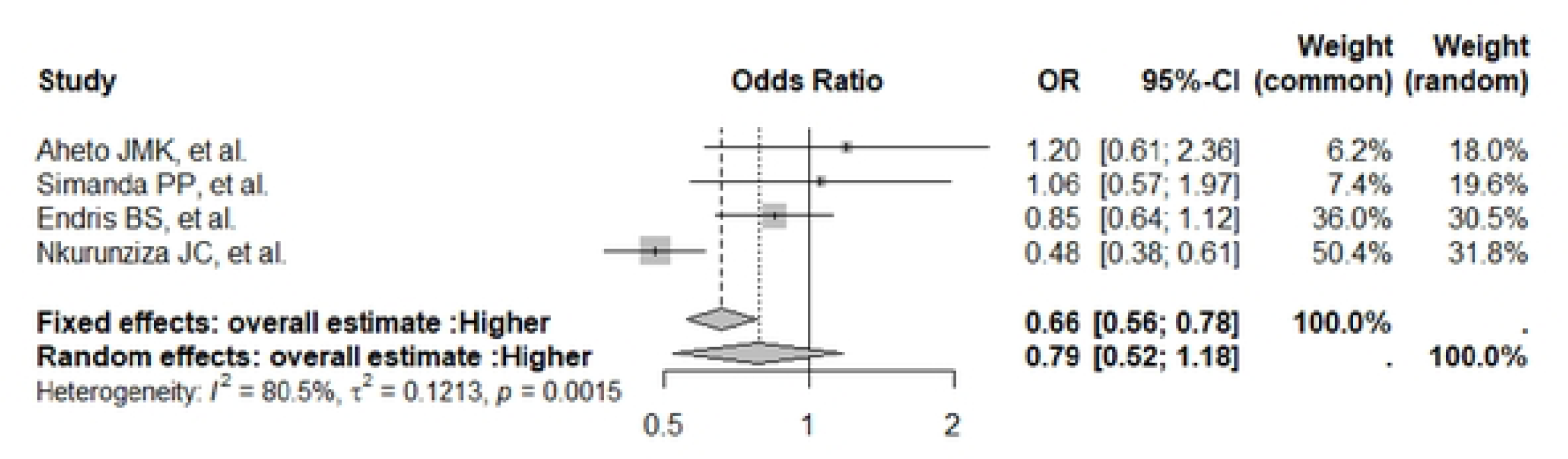
Mother education (higher).

**Figura 11:**
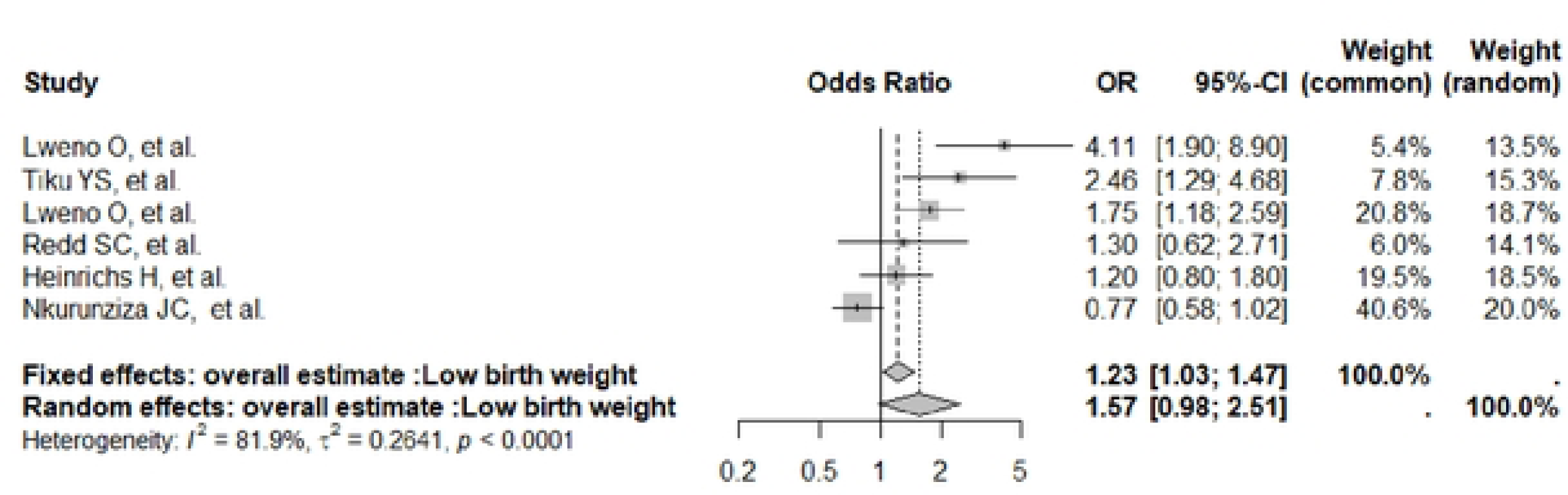
Child’s birth weight.

**Figura 12:**
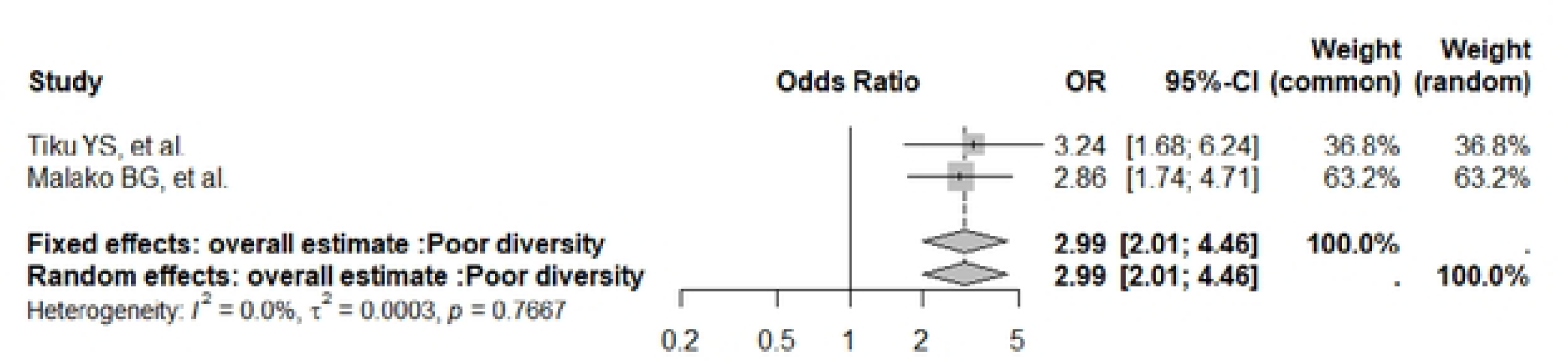
Child’s dietary diversity.

**Figura 13:**
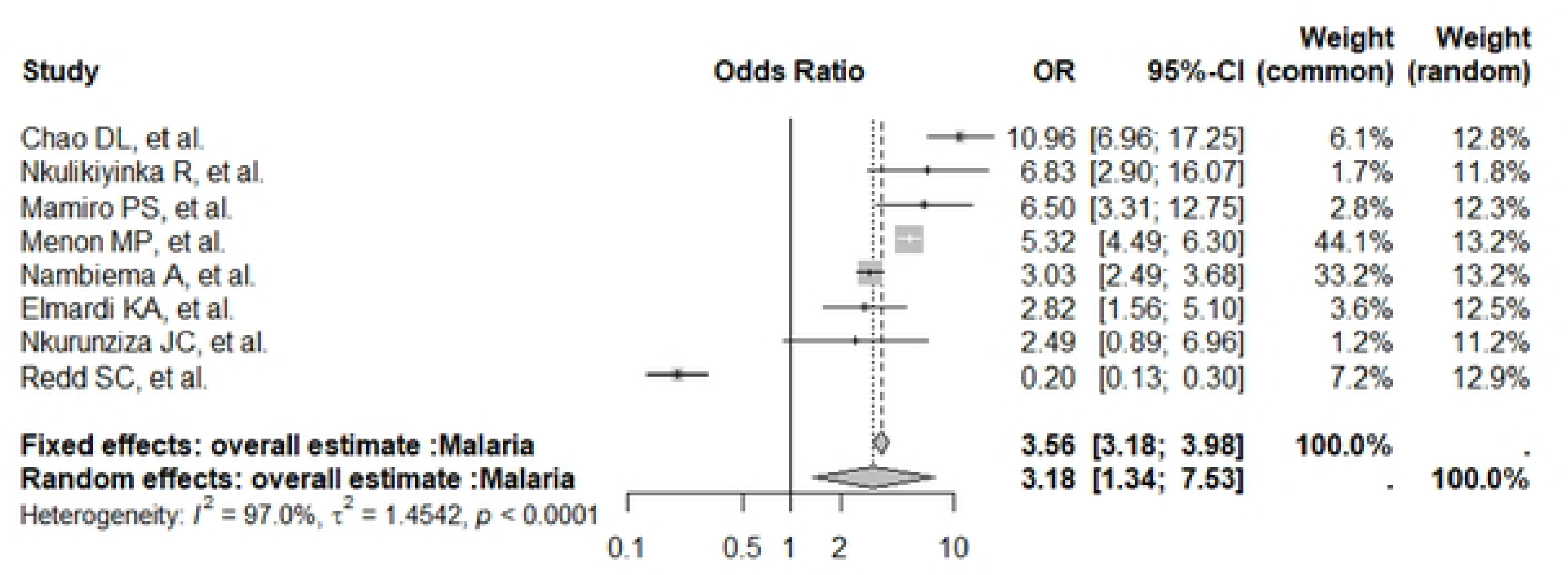
Malaria in the Child.

**Figura 14:**
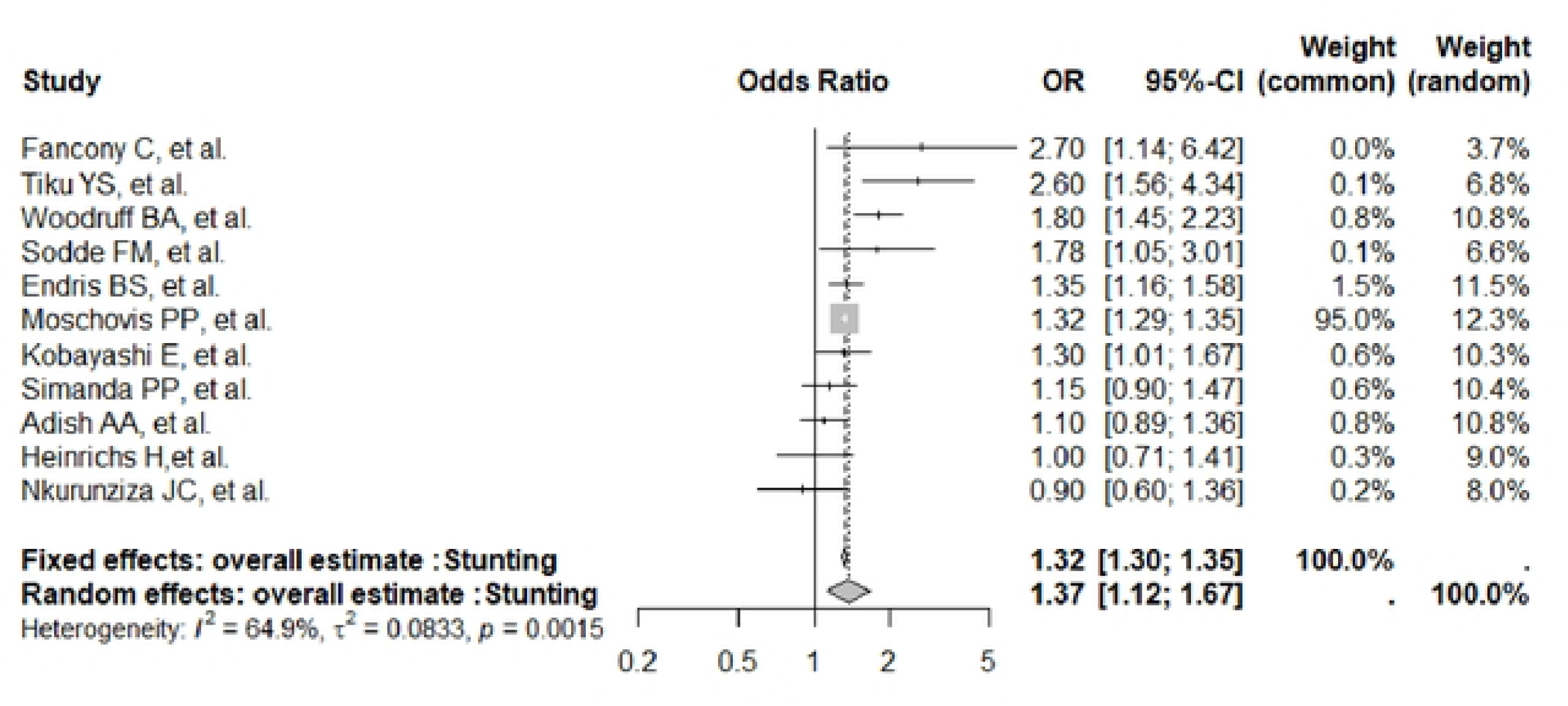
Stunting in the child.

**Figura 15:**
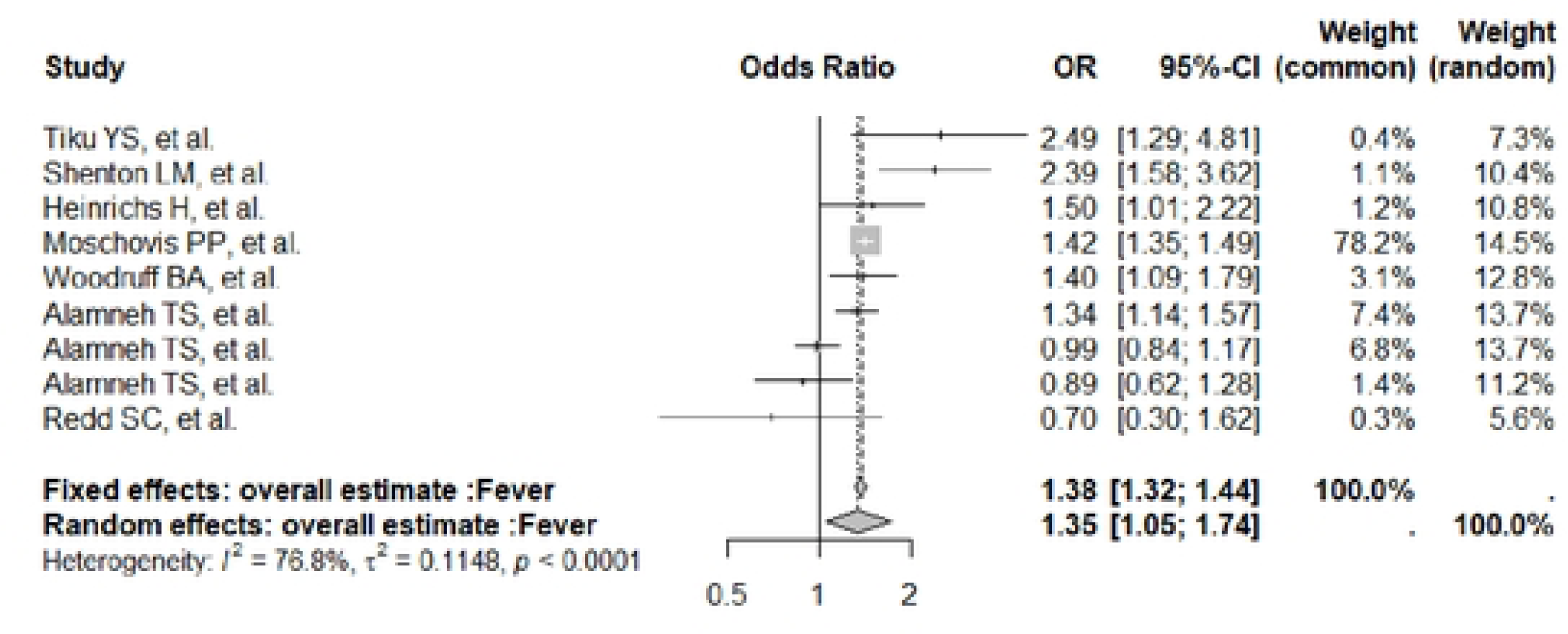
Fever in the child.

**Figura 16:**
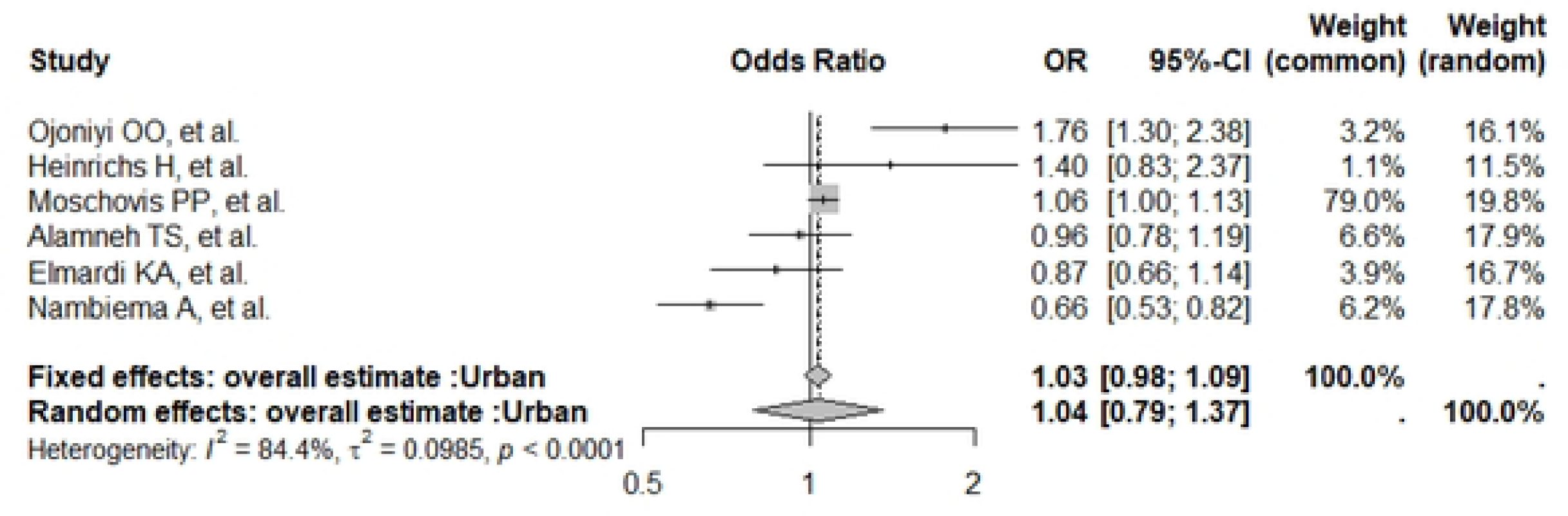
Urban residence.

**Figura 17:**
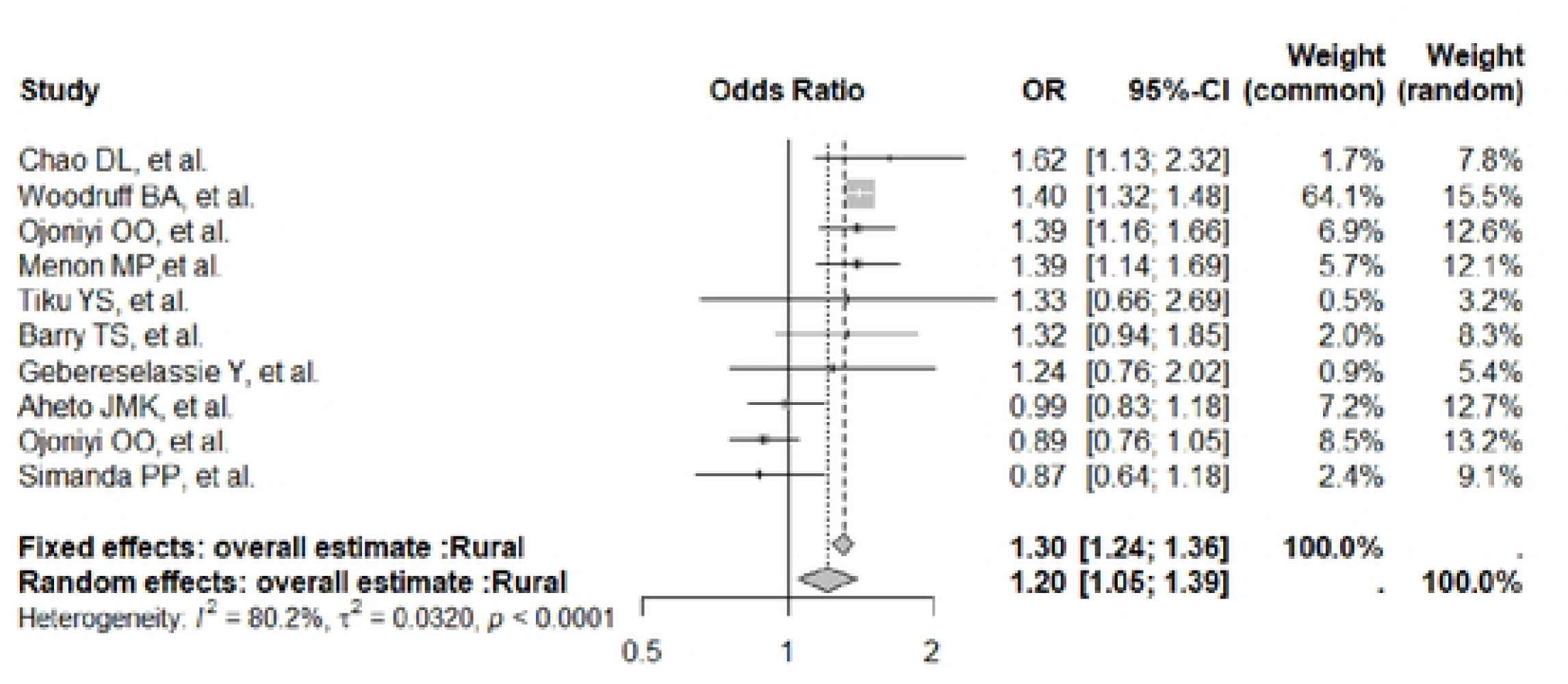
Rural residence.

**Figura 18:**
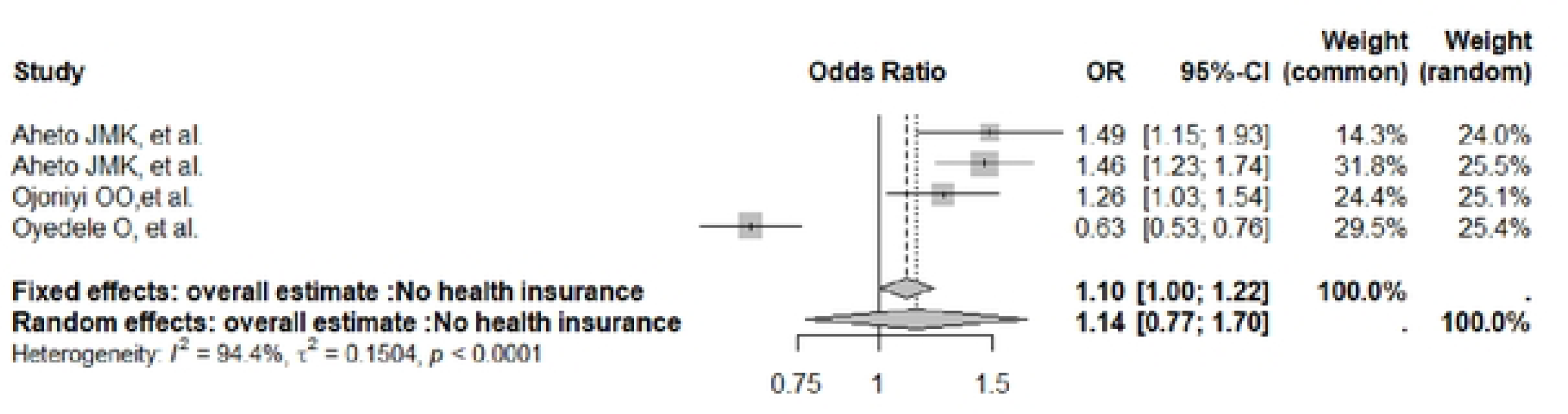
Health insurance.

**Figura 19:**
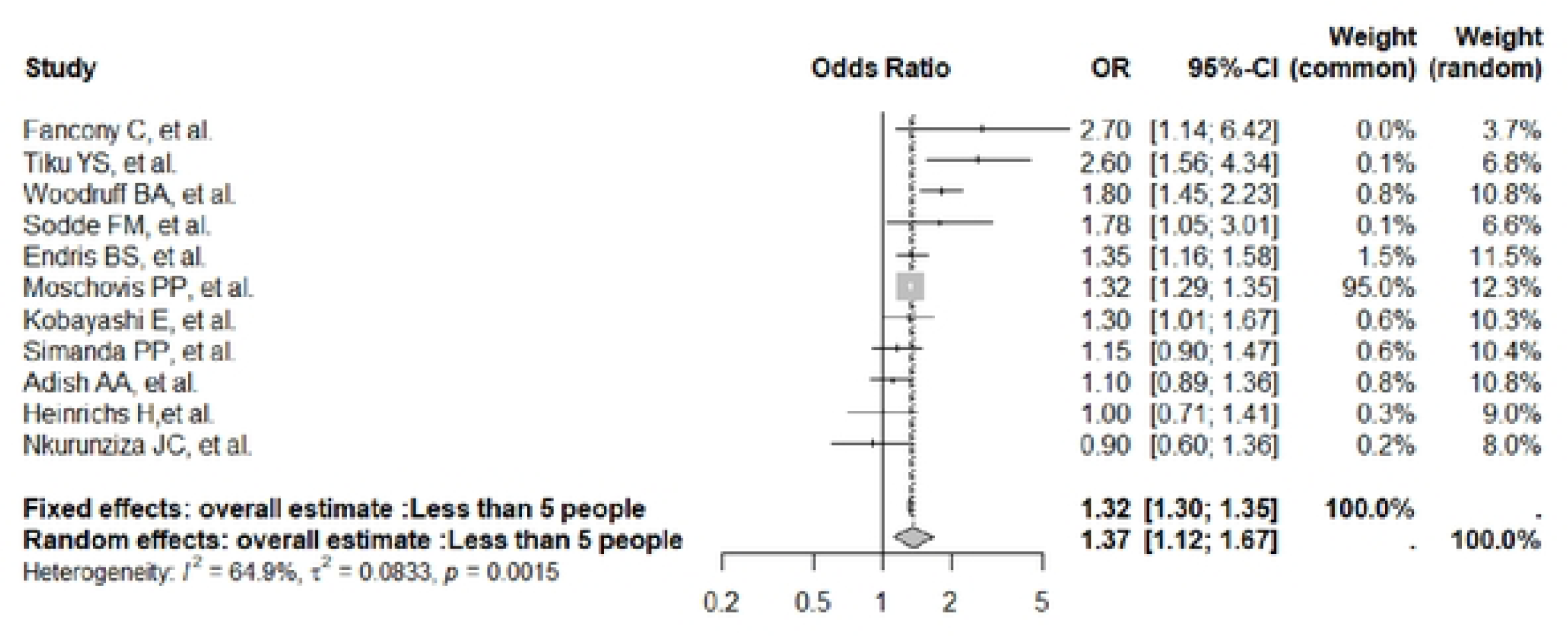
Family aggregate (less than 5).

**Figura 20:**
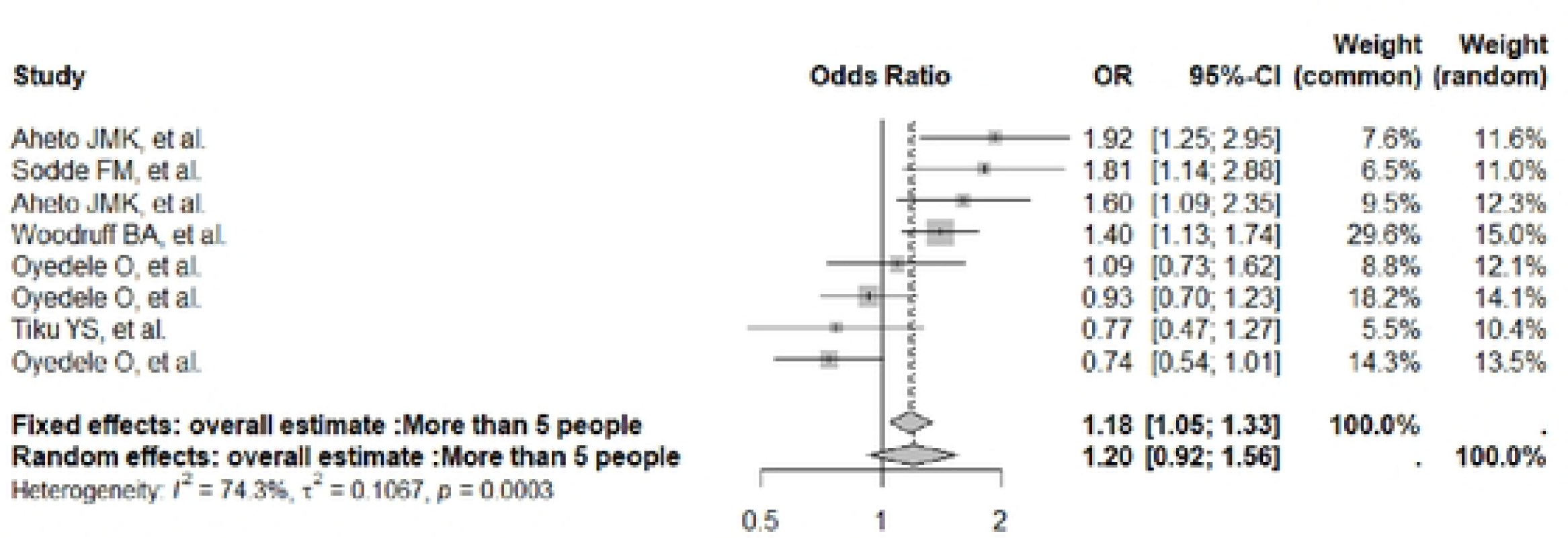
Family aggregate (1nore than 5).

**Figura 21:**
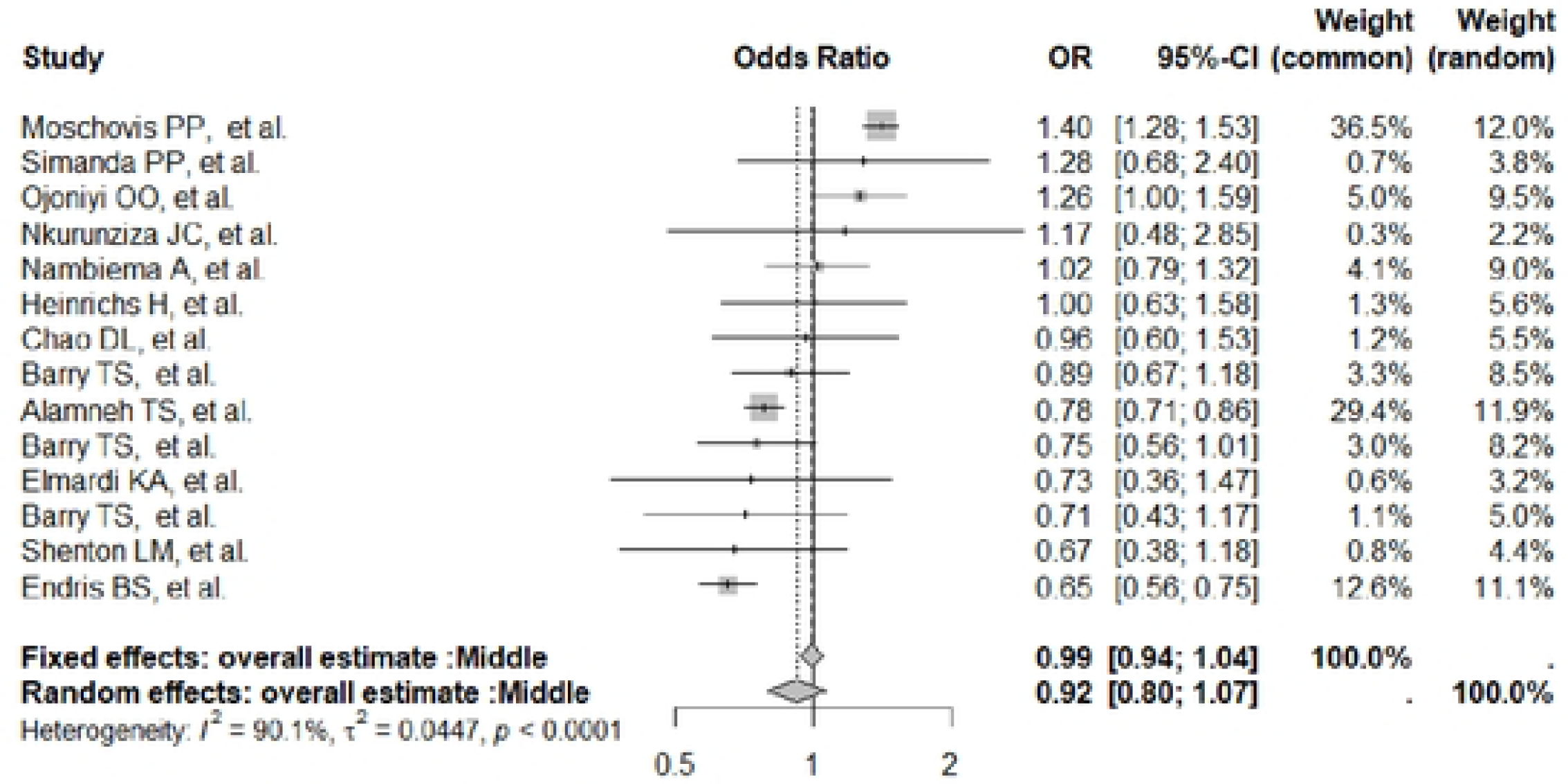
Econo1nic condition (n1iddle).

**Figura 22:**
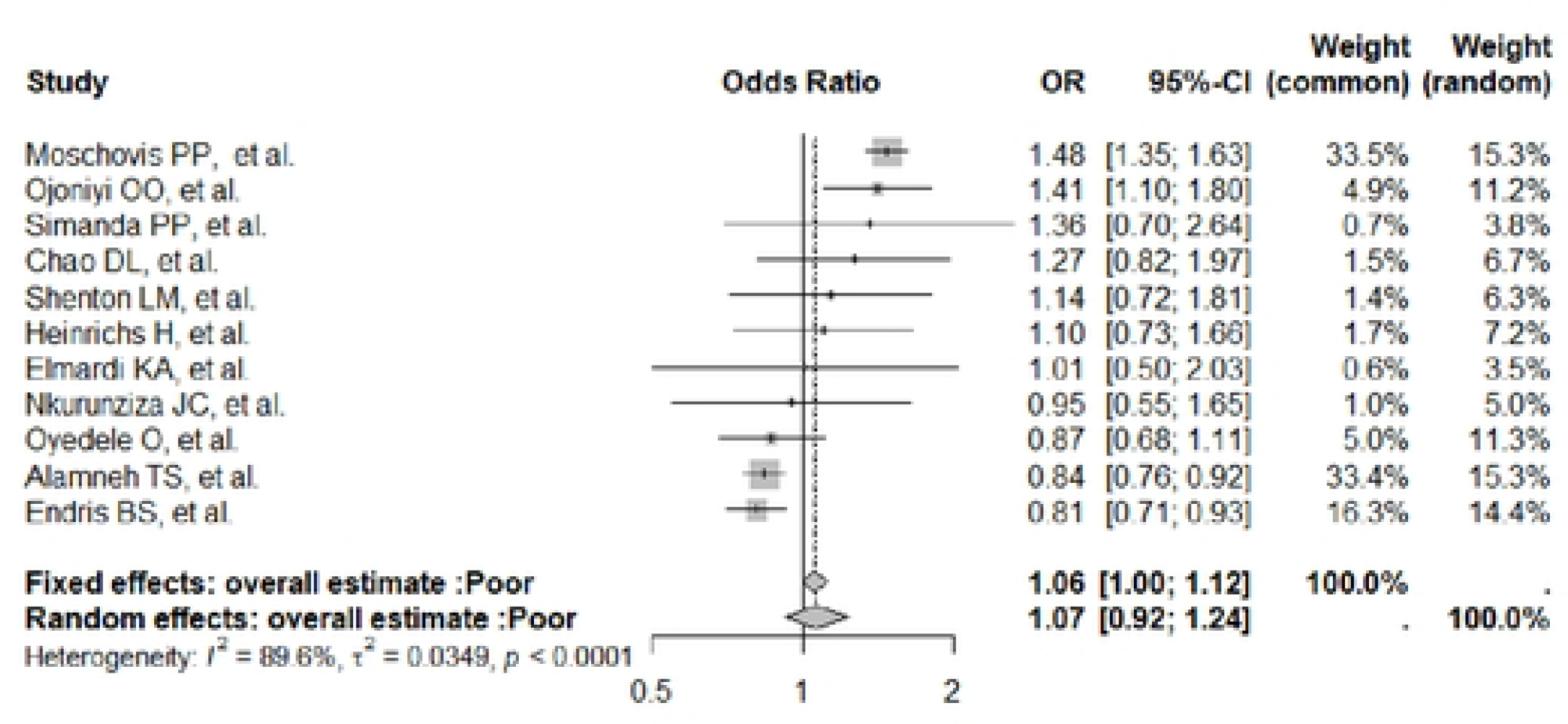
Econo111ic condition (poor).

**Figura 23:**
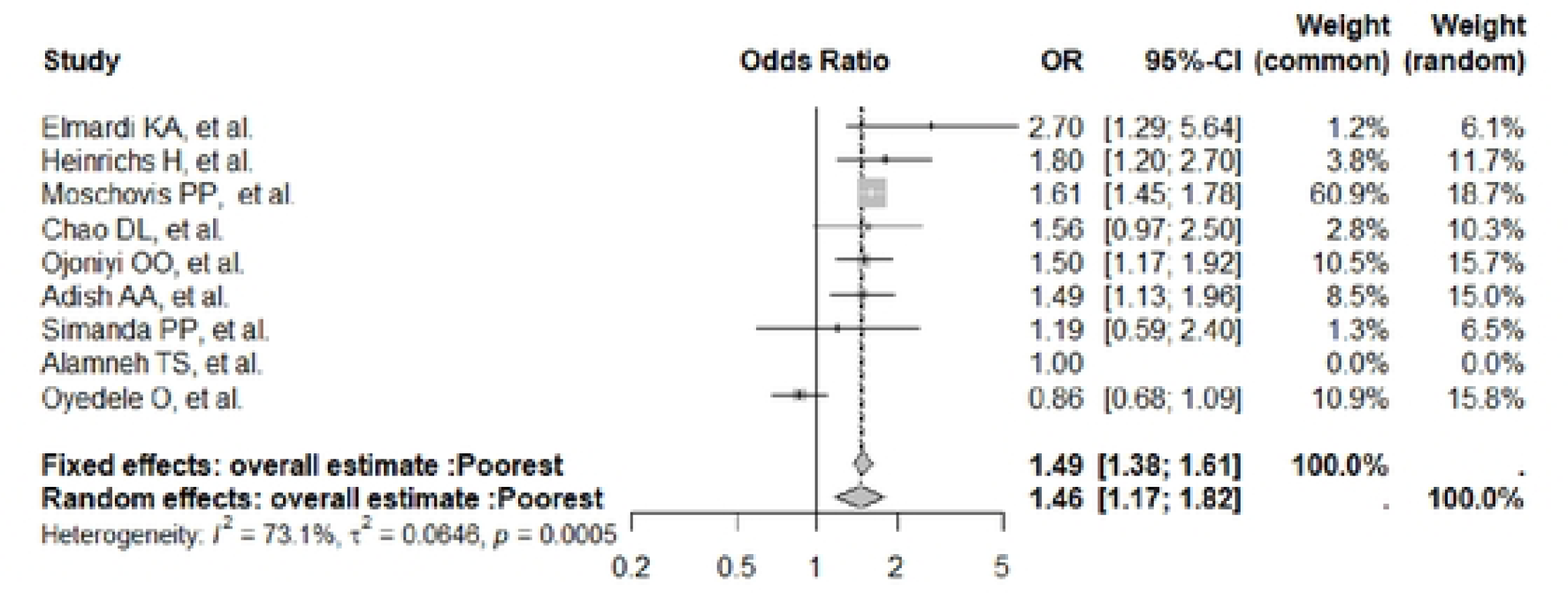
Economic condition (poorest).

**Figura 24:**
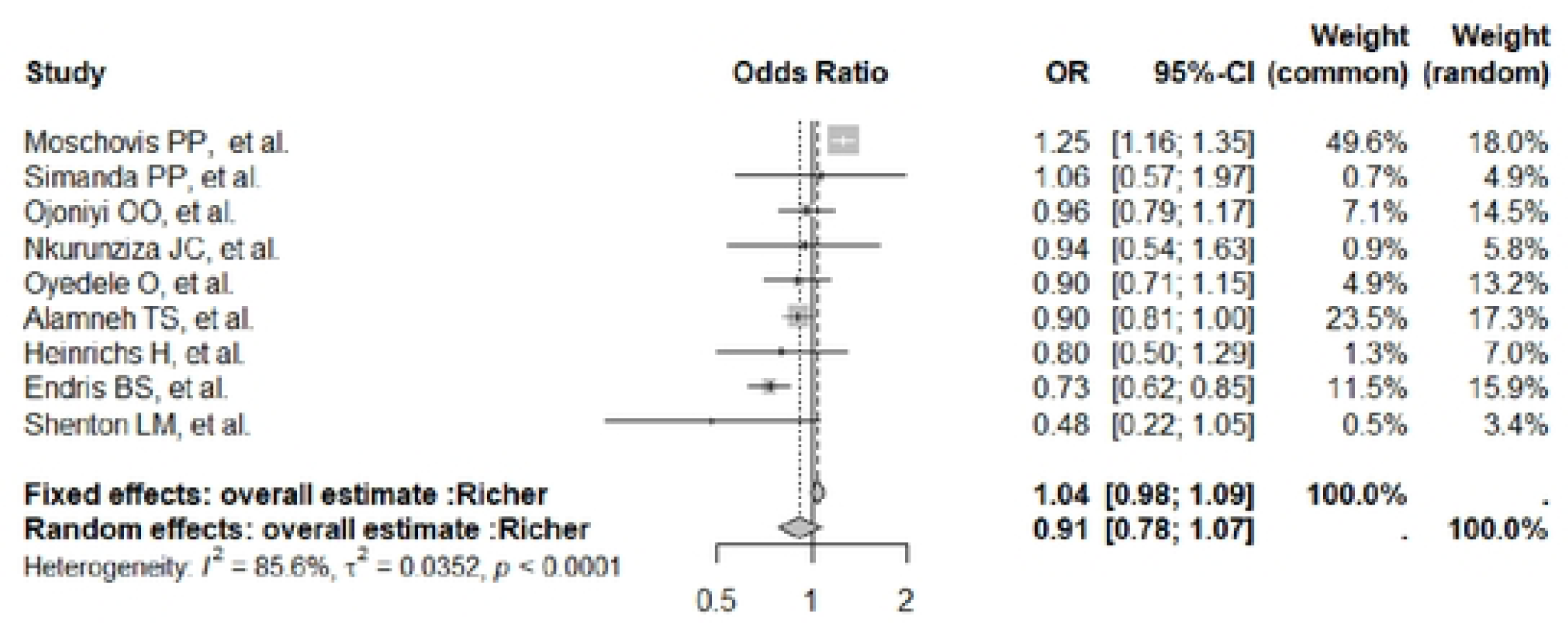
Economic condition (richer).

**Figura 25:**
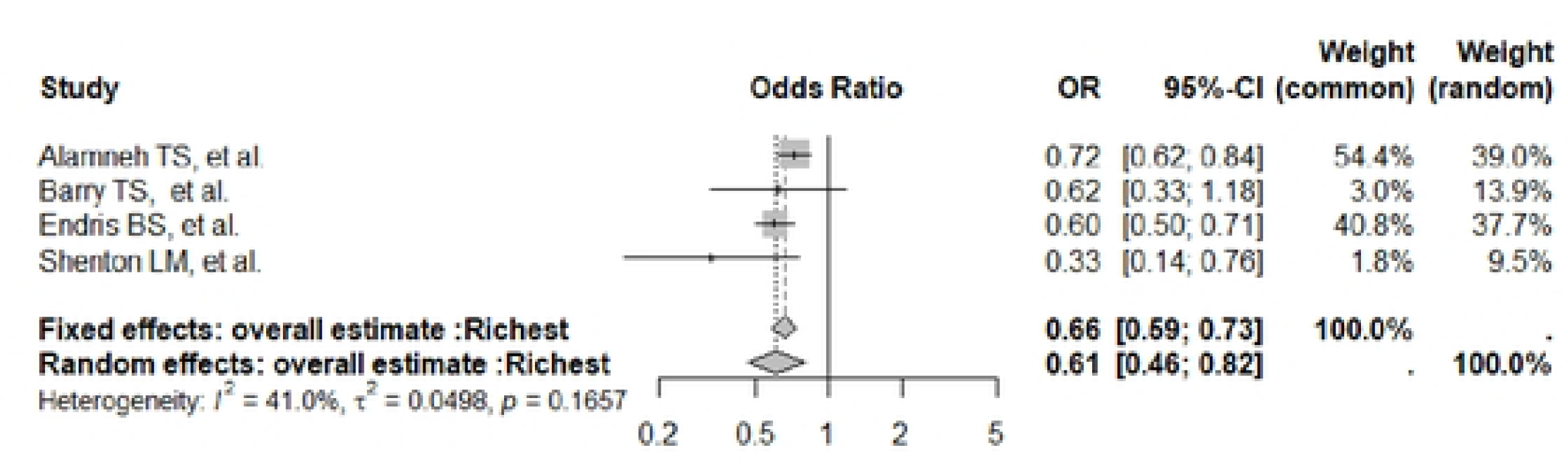
Economic condition (richest).

## References

1. Tadesse SE, Zerga AA, Mekonnen TC, Tadesse AW, Hussien FM, Feleke YW, et al. Burden and Determinants of Anemia among Under-Five Children in Africa: Systematic Review and Meta-Analysis. Anemia. Hindawi Limited; 2022. doi:10.1155/2022/1382940

2. World Health Organization. Haemoglobin concentrations for the diagnosis of anaemia and assessment of severity. 2011.

3. World Health Organization. Guideline on haemoglobin cutoffs to define anaemia in individuals and populations. 2024; 57.

4. McLean E, Cogswell M, Egli I, Wojdyla D, De Benoist B. Worldwide prevalence of anaemia, WHO Vitamin and Mineral Nutrition Information System, 1993-2005. Public Health Nutr. 2009;12: 444–454. doi:10.1017/S1368980008002401

5. Safiri S, Kolahi AA, Noori M, Nejadghaderi SA, Karamzad N, Bragazzi NL, et al. Burden of anemia and its underlying causes in 204 countries and territories, 1990–2019: results from the Global Burden of Disease Study 2019. J Hematol Oncol. 2021;14. doi:10.1186/s13045-021-01202-2

6. World Health Organization. Accelerating anaemia reduction A comprehensive framework for action. 2023.

7. Soares Magalhães RJ, Clements ACA. Spatial heterogeneity of haemoglobin concentration in preschool-age children in sub-Saharan Africa. Bull World Health Organ. 2011;89: 459–468. doi:10.2471/BLT.10.083568

8. Brunt DR, Grant CC, Wall CR, Reed PW. Interaction between risk factors for iron deficiency in young children. Nutrition and Dietetics. 2012;69: 285–292. doi:10.1111/j.1747-0080.2012.01597.x

9. Ngesa O, Mwambi H. Prevalence and risk factors of anaemia among children aged between 6 months and 14 years in Kenya. PLoS One. 2014;9. doi:10.1371/journal.pone.0113756

10. Egbi G, Steiner-Asiedu M, Kwesi FS aalia, Ayi I, Ofosu W, Setorglo J, et al. Anaemia among school children older than five years in the Volta Region of Ghana. Pan Afr Med J. 2014;17: 10. doi:10.11694/pamj.supp.2014.17.1.3205

11. Desalegn A, Mossie A, Gedefaw L. Nutritional iron deficiency anemia: Magnitude and its predictors among school age children, southwest ethiopia: A community based cross-sectional study. PLoS One. 2014;9. doi:10.1371/journal.pone.0114059

12. Seifu BL, Tesema GA. Individual-and community-level factors associated with anemia among children aged 6–23 months in sub-Saharan Africa: evidence from 32 sub-Saharan African countries. Archives of Public Health. 2022;80. doi:10.1186/s13690-022-00950-y

13. Belachew A, Tewabe T. Under-five anemia and its associated factors with dietary diversity, food security, stunted, and deworming in Ethiopia: Systematic review and meta-analysis. Systematic Reviews. BioMed Central Ltd.; 2020. doi:10.1186/s13643-020-01289-7

14. Gedfie S, Getawa S, Melku M. Prevalence and Associated Factors of Iron Deficiency and Iron Deficiency Anemia Among Under-5 Children: A Systematic Review and Meta-Analysis. Global Pediatric Health. SAGE Publications Inc.; 2022. doi:10.1177/2333794X221110860

15. Shamseer L, Moher D, Clarke M, Ghersi D, Liberati A, Petticrew M, et al. Preferred reporting items for systematic review and meta-analysis protocols (prisma-p) 2015: Elaboration and explanation. BMJ (Online). BMJ Publishing Group; 2015. doi:10.1136/bmj.g7647

16. van der Mierden S, Tsaioun K, Bleich A, Leenaars CHC. Software tools for literature screening in systematic reviews in biomedical research. Altex. ALTEX Edition; 2019. pp. 508–517. doi:10.14573/ALTEX.1902131

17. Tawfik GM, Dila KAS, Mohamed MYF, Tam DNH, Kien ND, Ahmed AM, et al. A step by step guide for conducting a systematic review and meta-analysis with simulation data. Tropical Medicine and Health. BioMed Central Ltd.; 2019. doi:10.1186/s41182-019-0165-6

18. Checklist for Systematic Reviews and Research Syntheses Critical Appraisal Checklist for Systematic Reviews and Research Syntheses 2. 2017. Available: http://joannabriggs.org/research/critical-appraisal-tools.html www.joannabriggs.org

19. Checklist for Analytical Cross Sectional Studies Critical Appraisal Checklist for Analytical Cross Sectional Studies 2. 2017. Available: http://joannabriggs.org/research/critical-appraisal-tools.html www.joannabriggs.org

20. Checklist for Cohort Studies. 2017. Available: http://joannabriggs.org/research/critical-appraisal-tools.html www.joannabriggs.org

21. Checklist for Case Control Studies. 2017. Available: http://joannabriggs.org/research/critical-appraisal-tools.html www.joannabriggs.org

22. Higgins JPT, Thompson SG, Deeks JJ, Altman DG. Measuring inconsistency in meta-analyses Testing for heterogeneity. 2002.

23. Higgins JPT, Thompson SG. Quantifying heterogeneity in a meta-analysis. Stat Med. 2002;21: 1539–1558. doi:10.1002/sim.1186

24. Egger M, Smith GD, Schneider M, Minder C. Papers Bias in meta-analysis detected by a simple, graphical test. 1997.

25. Begg CB, Mazumdar M. Operating Characteristics of a Rank Correlation Test for Publication Bias. 1994. Available: https://www.jstor.org/stable/2533446

26. Cooper HM., Hedges LV., Valentine JC. The handbook of research synthesis and meta-analysis. Russell Sage Foundation; 2012.

27. Silveira VNC, Carvalho CA, Viola PCAF, Magalhães EIS, Padilha LL, Conceição SIO, et al. Prevalence of iron-deficiency anaemia in Brazilian children under 5 years of age: A systematic review and meta-analysis. British Journal of Nutrition. Cambridge University Press; 2021. pp. 1257–1269. doi:10.1017/S000711452000522X

28. Stevens GA, Paciorek CJ, Flores-Urrutia MC, Borghi E, Namaste S, Wirth JP, et al. National, regional, and global estimates of anaemia by severity in women and children for 2000–19: a pooled analysis of population-representative data. Lancet Glob Health. 2022;10: e627–e639. doi:10.1016/S2214-109X(22)00084-5

29. Tesema GA, Worku MG, Tessema ZT, Teshale AB, Alem AZ, Yeshaw Y, et al. Prevalence and determinants of severity levels of anemia among children aged 6-59 months in sub-Saharan Africa: A multilevel ordinal logistic regression analysis. PLoS One. 2021;16. doi:10.1371/journal.pone.0249978

30. Liu Y, Ren W, Wang S, Xiang M, Zhang S, Zhang F. Global burden of anemia and cause among children under five years 1990–2019: findings from the global burden of disease study 2019. Front Nutr. 2024;11. doi:10.3389/fnut.2024.1474664

31. World Health Organization. Global Nutrition Targets 2025 Stunting Policy Brief.

32. Gedfie S, Getawa S, Melku M. Prevalence and Associated Factors of Iron Deficiency and Iron Deficiency Anemia Among Under-5 Children: A Systematic Review and Meta-Analysis. Global Pediatric Health. SAGE Publications Inc.; 2022. doi:10.1177/2333794X221110860

33. Habib MA, Black K, Soofi SB, Hussain I, Bhatti Z, Bhutta ZA, et al. Prevalence and predictors of iron deficiency anemia in children under five years of age in Pakistan, a secondary analysis of National Nutrition Survey data 2011-2012. PLoS One. 2016;11. doi:10.1371/journal.pone.0155051

34. Cohee LM, Opondo C, Clarke SE, Halliday KE, Cano J, Shipper AG, et al. Preventive malaria treatment among school-aged children in sub-Saharan Africa: a systematic review and meta-analyses. Lancet Glob Health. 2020;8: e1499– e1511. doi:10.1016/S2214-109X(20)30325-9

35. Nawa DN, Mpundu-Zimba P, Sialubanje C. Malaria Prevalence and Associated Additional Risk Factors among Children Under-Five Years Who Sleep under Insecticide Treated Nets in Zambia. Med J Zambia. 2024;50: 296–306. doi:10.55320/mjz.50.4.439

36. Trampuz A, Jereb M, Muzlovic I, Prabhu RM. Clinical review: Severe malaria. Critical Care. 2003. pp. 315–323. doi:10.1186/cc2183

37. Andrew M. Prentice HGCD and SEC. Iron metabolism and malaria, Food and Nutrition Bulletin, vol. 28, no. 4. 2007.

38. Preza GC, Pinon R, Ganz T, Nemeth E. Cellular Catabolism of the Iron-Regulatory Peptide Hormone Hepcidin. PLoS One. 2013;8. doi:10.1371/journal.pone.0058934

39. Milner EM, Kariger P, Pickering AJ, Stewart CP, Byrd K, Lin A, et al. Association between malaria infection and early childhood development mediated by anemia in rural Kenya. Int J Environ Res Public Health. 2020;17. doi:10.3390/ijerph17030902

40. Tadesse SE, Zerga AA, Mekonnen TC, Tadesse AW, Hussien FM, Feleke YW, et al. Burden and Determinants of Anemia among Under-Five Children in Africa: Systematic Review and Meta-Analysis. Anemia. Hindawi Limited; 2022. doi:10.1155/2022/1382940

41. Malako BG, Asamoah BO, Tadesse M, Hussen R, Gebre MT. Stunting and anemia among children 6-23 months old in Damot Sore district, Southern Ethiopia. BMC Nutr. 2019;5. doi:10.1186/s40795-018-0268-1

42. Gosdin L, Martorell R, Bartolini RM, Mehta R, Srikantiah S, Young MF. The co-occurrence of anaemia and stunting in young children. Matern Child Nutr. 2018;14. doi:10.1111/mcn.12597

43. Silla LM da R, Zelmanowicz A, Mito I, Michalowski M, Hellwing T, Shilling MA, et al. High Prevalence of Anemia in Children and Adult Women in an Urban Population in Southern Brazil. PLoS One. 2013;8. doi:10.1371/journal.pone.0068805

44. Gebreegziabher E, Bountogo M, Sié A, Zakane A, Compaoré G, Ouedraogo T, et al. Influence of maternal age on birth and infant outcomes at 6 months: a cohort study with quantitative bias analysis. Int J Epidemiol. 2023;52: 414–425. doi:10.1093/ije/dyac236

45. Fall CHD, Sachdev HS, Osmond C, Restrepo-Mendez MC, Victora C, Martorell R, et al. Association between maternal age at childbirth and child and adult outcomes in the offspring: A prospective study in five low-income and middle-income countries (COHORTS collaboration). Lancet Glob Health. 2015;3: e366– e377. doi:10.1016/S2214-109X(15)00038-8

46. Obasohan PE, Walters SJ, Jacques R, Khatab K. A scoping review of the risk factors associated with anaemia among children under five years in sub-Saharan African countries. International Journal of Environmental Research and Public Health. MDPI AG; 2020. pp. 1–20. doi:10.3390/ijerph17238829

47. Moschovis PP, Wiens MO, Arlington L, Antsygina O, Hayden D, Dzik W, et al. Individual, maternal and household risk factors for anaemia among young children in sub-Saharan Africa: A cross-sectional study. BMJ Open. 2018;8. doi:10.1136/bmjopen-2017-019654

48. Kebede TB, Mengesha S, Lindtjorn B, Engebretsen IMS. Anaemia, anthropometric undernutrition and associated factors among mothers with children younger than 2 years of age in the rural Dale district, southern Ethiopia: A community-based study. Matern Child Nutr. 2022;18. doi:10.1111/mcn.13423

49. Nambiema A, Robert A, Yaya I. Prevalence and risk factors of anemia in children aged from 6 to. BMC Public Health. 2019;19. doi:10.1186/s12889-019-6547-1

50. Shibeshi AH, Mare KU, Kase BF, Wubshet BZ, Tebeje TM, Asgedom YS, et al. The effect of dietary diversity on anemia levels among children 6-23 months in sub-Saharan Africa: A multilevel ordinal logistic regression model. PLoS One. 2024;19. doi:10.1371/journal.pone.0298647

51. Visser M, Van Zyl T, Hanekom SM, Baumgartner J, van der Hoeven M, Taljaard-Krugell C, et al. Nutrient patterns and their relation to anemia and iron status in 5-to 12-y-old children in South Africa. Nutrition. 2019;62: 194–200. doi:10.1016/j.nut.2019.01.016

52. Gebreweld A, Ali N, Ali R, Fisha T. Prevalence of anemia and its associated factors among children under five years of age attending at Guguftu health center, South Wollo, Northeast Ethiopia. PLoS One. 2019;14. doi:10.1371/journal.pone.0218961

53. Mbunga BK, Mapatano MA, Strand TA, Gjengedal ELF, Akilimali PZ, Engebretsen IMS. Prevalence of anemia, iron-deficiency anemia, and associated factors among children aged 1–5 years in the rural, malaria-endemic setting of popokabaka, democratic Republic of Congo: A cross-sectional study. Nutrients. 2021;13: 1–13. doi:10.3390/nu13031010

54. Turawa E, Awotiwon O, Dhansay MA, Cois A, Labadarios D, Bradshaw D, et al. Prevalence of anaemia, iron deficiency, and iron deficiency anaemia in women of reproductive age and children under 5 years of age in south africa (1997–2021): A systematic review. Int J Environ Res Public Health. 2021;18. doi:10.3390/ijerph182312799

55. Chao DL, Oron AP, Chabot-Couture G, Sopekan A, Nnebe-Agumadu U, Bates I, et al. Contribution of malaria and sickle cell disease to anaemia among children aged 6-59 months in Nigeria: a cross-sectional study using data from the 2018 Demographic and Health Survey. BMJ Open. 2022;12. doi:10.1136/bmjopen-2022-063369

56. Premji Z, Hamisi Y, Shift C, Minjas L’z J, Lubega P, Makwaya C. Anaemia and Plasmodium falciparum infections among young children in an holoendemic area, Bagamoyo, Tanzania. Acta Trop. 1995.

57. Tiku YS, Mekonnen TC, Workie SB, Amare E. Does anaemia have major public health importance in children aged 6-59 months in the duggina fanigo district of wolaita zone, Southern Ethiopia? Ann Nutr Metab. 2018;72: 3–11. doi:10.1159/000484324

58. Gebereselassie Y, Birhanselassie M, Menjetta T, Alemu J, Tsegaye A. Magnitude, Severity, and Associated Factors of Anemia among Under-Five Children Attending Hawassa University Teaching and Referral Hospital, Hawassa, Southern Ethiopia, 2016. Anemia. 2020;2020. doi:10.1155/2020/7580104

59. Kobayashi E, Negi B, Nakazawa M. The association between food groups and childhood anemia in Zambia, based on the analysis of Zambia Demographic and Health Survey 2018. J Public Health Afr. 2022;13. doi:10.4081/jphia.2022.2199

60. Shimanda PP, Amukugo HJ, Norström F. Socioeconomic factors associated with anaemia among children aged 6–59 months in Namibia. J Public Health Afr. 2020;11. doi:10.4081/jphia.2020.1131

61. Donahue Angel M, Berti P, Siekmans K, Tugirimana PL, Boy E. Prevalence of Iron Deficiency and Iron Deficiency Anemia in the Northern and Southern Provinces of Rwanda. Food Nutr Bull. 2017;38: 554–563. doi:10.1177/0379572117723134

62. Aheto JMK, Alhassan Y, Puplampu AE, Boglo JK, Sedzro KM. Anemia prevalence and its predictors among children under-five years in Ghana. A multilevel analysis of the cross-sectional 2019 Ghana Malaria Indicator Survey. Health Sci Rep. 2023;6. doi:10.1002/hsr2.1643

63. Sodde FM, Liga AD, Jabir YN, Tamiru D, Kidane R. Magnitude and predictors of anemia among preschool children (36–59 months) in Atingo town, Jimma, Ethiopia. Health Sci Rep. 2023;6. doi:10.1002/hsr2.1358

64. Barry TS, Ngesa O, Onyango NO, Mwambi H. Bayesian spatial modeling of anemia among children under 5 years in guinea. Int J Environ Res Public Health. 2021;18. doi:10.3390/ijerph18126447

65. Kajoba D, Egesa WI, Muyombya S, Ortiz YA, Nduwimana M, Ndeezi G. Prevalence and Factors Associated with Iron Deficiency Anaemia among Children Aged 6-23 Months in Southwestern Uganda. International Journal of Pediatrics (United Kingdom). 2024;2024. doi:10.1155/2024/6663774

66. Nkulikiyinka R, Binagwaho A, Palmer K. The changing importance of key factors associated with anaemia in 6-to 59-month-old children in a sub-Saharan African setting where malaria is on the decline: Analysis of the Rwanda Demographic and Health Survey 2010. Tropical Medicine and International Health. 2015;20: 1722–1732. doi:10.1111/tmi.12604

67. Lweno O, Hertzmark E, Darling AM, Noor R, Bakari L, Sudfeld C, et al. The High Burden and Predictors of Anemia Among Infants Aged 6 to 12 Months in Dar es Salaam, Tanzania. Food Nutr Bull. 2022;43: 68–83. doi:10.1177/03795721211007009

68. Mamiro PS, Kolsteren P, Roberfroid D, Tatala S, Opsomer AS, Van Camp JH. Feeding Practices and Factors Contributing to Wasting, Stunting, and Iron-deficiency Anaemia among 3–23-month Old Children in Kilosa District, Rural Tanzania. Source: Journal of Health. 2005. Available: https://www.jstor.org/stable/23499322

69. Heinrichs H, Endris BS, Dejene T, Dinant GJ, Spigt M. Anaemia and its determinants among young children aged 6–23 months in Ethiopia (2005–2016). Matern Child Nutr. 2021;17. doi:10.1111/mcn.13082

70. Oyedele O. Childhood anaemia levels among under-5 children in Namibia and their associated sociodemographic factors: A multivariate ordinal modelling approach. Nutr Health. 2024;30: 573–586. doi:10.1177/02601060221129695

71. Kejo D, Petrucka P, Martin H, Kimanya M, Mosha T. Prevalence and predictors of anemia among children under 5 years of age in Arusha District, Tanzania. Pediatric Health Med Ther. 2018;Volume 9: 9–15. doi:10.2147/phmt.s148515

72. Adish AA, Esrey SA, Gyorkos TW, Johns T. Risk factors for iron deficiency anaemia in preschool children in northern Ethiopia. Public Health Nutr. 1999;2: 243–252. doi:10.1017/s1368980099000336

73. Shenton LM, Jones AD, Wilson ML. Factors Associated with Anemia Status Among Children Aged 6–59 months in Ghana, 2003–2014. Matern Child Health J. 2020;24: 483–502. doi:10.1007/s10995-019-02865-7

74. Malako BG, Teshome MS, Belachew T. Anemia and associated factors among children aged 6-23 months in Damot Sore District, Wolaita Zone, South Ethiopia. BMC Hematol. 2018;18. doi:10.1186/s12878-018-0108-1

75. Ojoniyi OO, Odimegwu CO, Olamijuwon EO, Akinyemi JO. Does education offset the effect of maternal disadvantage on childhood anaemia in Tanzania? Evidence from a nationally representative cross-sectional study. BMC Pediatr. 2019;19. doi:10.1186/s12887-019-1465-z

76. Fançony C, Soares Â, Lavinha J, Barros H, Brito M. Iron deficiency anaemia among 6-to-36-month children from northern Angola. BMC Pediatr. 2020;20. doi:10.1186/s12887-020-02185-8

77. Elmardi KA, Adam I, Malik EM, Ibrahim AA, Elhassan AH, Kafy HT, et al. Anaemia prevalence and determinants in under 5 years children: findings of a cross-sectional population-based study in Sudan. BMC Pediatr. 2020;20. doi:10.1186/s12887-020-02434-w

78. Menon MP, Yoon SS. Prevalence and factors associated with anemia among children under 5 years of age-Uganda, 2009. American Journal of Tropical Medicine and Hygiene. 2015;93: 521–526. doi:10.4269/ajtmh.15-0102

79. Endris BS, Dinant GJ, Gebreyesus SH, Spigt M. Risk factors of anemia among preschool children in Ethiopia: a Bayesian geo-statistical model. BMC Nutr. 2022;8. doi:10.1186/s40795-021-00495-3

80. Alamneh TS, Melesse AW, Gelaye KA. Determinants of anemia severity levels among children aged 6–59 months in Ethiopia: Multilevel Bayesian statistical approach. Sci Rep. 2023;13. doi:10.1038/s41598-022-20381-7

81. Redd SC, Wirima JJ, Steketee RW. RISK FACTORS FOR ANEMIA IN YOUNG CHILDREN IN RURAL MALAWI. Am J Trop Med itig. 1994.

82. Kahigwa E, Schellenberg D, Sanz S, Aponte JJ, Wigayi J, Mshinda H, et al. Risk factors for presentation to hospital with severe anaemia in Tanzanian children: A case-control study. Tropical Medicine and International Health. 2002;7: 823–830. doi:10.1046/j.1365-3156.2002.00938.x

83. Woodruff BA, Wirth JP, Ngnie-Teta I, Beaulière JM, Mamady D, Ayoya MA, et al. Determinants of Stunting, Wasting, and Anemia in Guinean Preschool-Age Children: An Analysis of DHS Data From 1999, 2005, and 2012. Food Nutr Bull. 2018;39: 39–53. doi:10.1177/0379572117743004

